# Immune Biomarker Signatures as Predictors of Functional and Pain Recovery After Total Knee Arthroplasty in Older Adults

**DOI:** 10.64898/2026.06.08.26355189

**Authors:** Virginia Byers Kraus, Noah D Greenberg, Marissa Ashner, Janet L Huebner, Akshay Bareja, Sarah Peskoe, Corey Simon, Heather E Whitson, Cathleen S Colón-Emeric

## Abstract

Postoperative resilience varies widely among older adults, yet the biological drivers of recovery remain unclear. We evaluated whether preoperative immune profiles—measured in plasma and through *ex vivo* whole-blood stimulation—predict resilience to the acute stress of total knee arthroplasty. A total of 152 adults (≥60 years) in the PRIME-KNEE cohort underwent elective total knee arthroplasty and had available blood samples for measurement of 45 immune biomarkers, quantified in plasma and in whole blood stimulated *ex vivo* for 24 hours with lipopolysaccharide (LPS) or influenza antigen (FLU). Resilience was assessed using Expected Recovery Differential (ERD) and Resilience Trajectory (RT) across pain severity, pain interference, lower-extremity physical activities of daily living (LE-PADLs), and step counts. An exploratory stability-selection framework using LASSO identified biomarker predictors of postoperative outcomes. Plasma and stimulated biomarkers showed broadly similar predictive performance. A shared set of biomarkers—including LBP, leptin, TNFR1, CD30, and LIF—was consistently selected across models. Immune predictors explained ∼12-24% of the variance in resilience outcomes. Distinct immune signatures emerged for pain versus functional recovery: pain-related predictors mapped to local inflammatory and neuroimmune pathways, whereas function-related predictors reflected systemic inflammatory load and cytokine signaling. Preoperative immune biomarkers, whether measured in plasma or after *ex vivo* stimulation, capture meaningful variance in postoperative resilience. The divergence between pain-related and function-related immune signatures highlights biologically distinct pathways underlying different dimensions of recovery and supports further development of immune-based perioperative risk assessment.

**Author Summary:** In this study, we explored why recovery after knee replacement surgery varies so widely among older adults. While many people regain mobility and experience pain relief, others continue to struggle with discomfort and limited function. We asked whether differences in the immune system before surgery might help explain these outcomes. To do this, we measured a wide range of immune signals in blood samples collected before surgery and then followed participants over several months, tracking their pain, daily activities, and physical movement. We found that patterns in these immune signals were linked to how well people recovered. Importantly, some of the signals that predicted pain recovery were different from those linked to improvements in physical function, suggesting that these aspects of recovery are driven by distinct biological processes. Our findings place the immune system at the center of recovery from major surgery and highlight its potential as a tool for identifying who may need more support. In the future, this knowledge could help guide more personalized approaches to care and improve recovery for older adults undergoing surgery.

**Graphical Abstract:** Overview of the PRIME-KNEE Study Workflow and Immune Signatures Predicting Postoperative Resilience.
Pre-operative blood was collected from older adults undergoing elective total knee arthroplasty. Whole blood was incubated under three conditions—unstimulated (NULL), lipopolysaccharide (LPS), or influenza antigen (FLU)—using the TruCulture® system to assess dynamic immune responsiveness. Forty-five pro- and anti-inflammatory biomarkers were quantified in plasma and stimulated culture supernatants. Postoperative pain, pain interference, lower-extremity physical activities of daily living (LE-PADLs), and step counts were used to derive two resilience metrics: Expected Recovery Differential (ERD) and Resilience Trajectory (RT). Stability-selection LASSO models identified pre-operative immune predictors of postoperative resilience. Panels A and B illustrate distinct immune signatures associated with pain versus functional outcomes. **Panel A:** Pain-related predictors (IFN-α2a, IL-15, IL-1Ra, LBP, LIF, SAA) cluster around local inflammatory and neuroimmune pathways implicated in nociceptor sensitization. **Panel B:** Function-related predictors (CRP, eotaxin, gp130, IL-6) reflect systemic inflammatory load and broad cytokine signaling associated with reduced functional capacity. Together, these findings highlight biologically distinct immune pathways underlying pain and functional recovery after knee arthroplasty. Graphic created in BioRender.

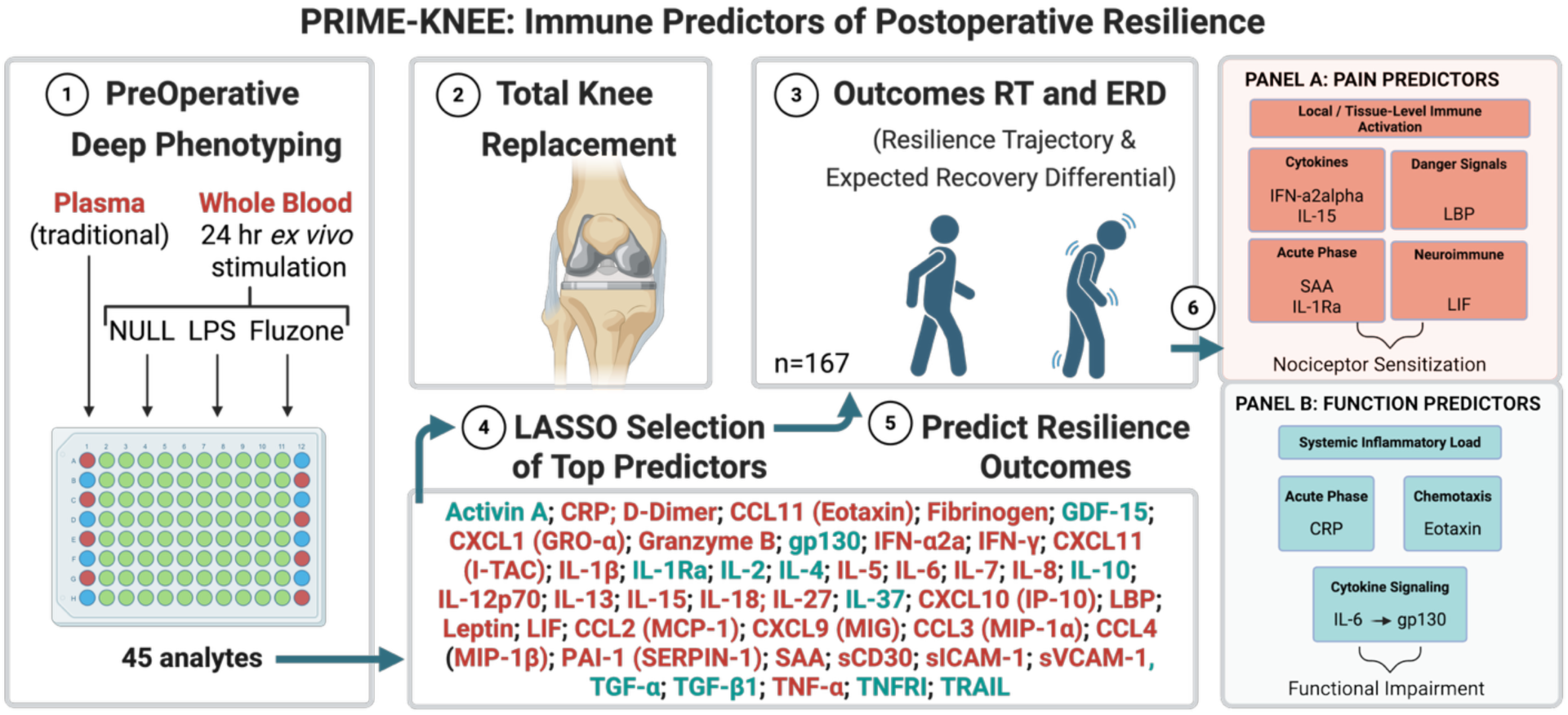

## Introduction

Resilience—the capacity to recover or maintain function in response to physiological stressors—is a critical determinant of health outcomes in older adults. At the molecular and cellular levels, resilience is shaped by complex biological interactions that remain incompletely understood. A dynamic systems approach, in which controlled perturbations are used to assess physiological responses, has been employed to identify subsystems and pathways involved in resilience. Such “biomarker stress tests” are modeled after cardiac stress tests and can be performed *in vivo* (e.g., serum cartilage oligomeric matrix protein mechanosensitivity, Adrenocorticotropic Hormone [ACTH] stimulation, glucose tolerance testing) (1–3) or *ex vivo* (whole blood immune cell reactivity to stimulants (4) such as lipopolysaccharide [LPS] (5, 6) or influenza vaccine antigen (7)) with the underlying expectation that *ex vivo* responses will reflect *in vivo* biological resilience.

The *ex vivo* whole blood LPS stimulation assay has been widely used to explore individual cytokine reactivity as part of human innate immune responses (8). Interestingly, both hypo- and hyperreactivity of immune analytes can be detrimental: hyporeactivity in sepsis (TNFα) (9) or COVID-19 (IL-1b, IL-12, IL-17a) (10), and hyperreactivity in COVID-19 (IL-6) (11). In osteoarthritis (OA), an innate immune profile characterized by elevated *ex vivo* LPS-stimulated IL-1β and IL-1Ra and reduced IL-10 production is associated with increased disease risk (5). Likewise, *ex vivo* whole blood influenza antigen stimulation has been used to profile vaccine-induced cytokine signatures (12).

Dynamic (stimulated) biomarkers may yield insights that synergize with static measures to more fully characterize biological responses. However, several questions remain unanswered: which biomarkers best predict resilient outcomes; which stimulation paradigms are most effective; and do absolute, relative, or ratio-based value parameterizations provide the most useful information for predicting resilient outcomes? To address these gaps, we applied multiple analytic strategies to evaluate plasma (unstimulated) and *ex vivo* stimulated immune responses and their association with resilience after surgery. Specifically, for *ex vivo* stimulated responses we modeled cytokine concentrations as (1) absolute stimulated values, (2) ratios of stimulated to unstimulated (null) values, and (3) percentage changes from null values—each compared against traditional plasma values of the same biomarkers.

Our overall objective was to gain a better understanding of the role of human immune function in resilience outcomes following acute surgical stress. We addressed this objective using the Physical Resilience Indicators and Mechanisms in the Elderly (PRIME) Knee Arthroplasty (PRIME-KNEE) study, a longitudinal observational cohort study of older adults undergoing total knee arthroplasty (TKA) (13). PRIME-KNEE quantified resilience using two approaches—the Expected Recovery Differential (ERD) and Resilience Trajectory (RT) phenotypes. ERD compares observed recovery with individualized recovery predictions based on multiple baseline clinical characteristics. RT models recovery patterns using serial outcome measures obtained after the stressor (14, 15).

To assess immune reactivity, we employed the TruCulture® (TC) platform, a standardized *ex vivo* whole blood stimulation system that yields cytokine concentrations under basal unstimulated (NULL) or stimulated (with LPS or Fluzone® [FLU]) conditions. This approach preserves physiologic cellular interactions and enables reproducible assessment of dynamic immune responses, which we compared to static plasma cytokine measures obtained from matched samples.

A key technical challenge—variation in sample dilution inherent to TC collection—was addressed using urea as an internal normalization marker. Because urea is neither synthesized nor metabolized in the *ex vivo* whole blood setting, comparison of matched plasma and TC urea levels enabled correction for dilution effects as we previously applied in other contexts (16), improving the accuracy of cytokine quantification.

Ultimately, this study sought to improve prediction of post-surgical resilience by identifying biologically meaningful markers of immune capacity. We hypothesized that preoperative immune profiles would predict resilience to the acute stress of knee arthroplasty.

## Results

### Cohort Description

Of the 203 enrolled participants, 199 completed surgery and 183 completed follow-up through 6 months post-surgery. A total of 152 of the 183 had available baseline biomarkers; this cohort is described in **Table 1**. The average age of the cohort was 71 years, the majority of whom identified as women (60.5%) and white (82.6%). The cohort was categorized as obese based on the average BMI of 31.6 kg/m^2^.

**Table 1.**
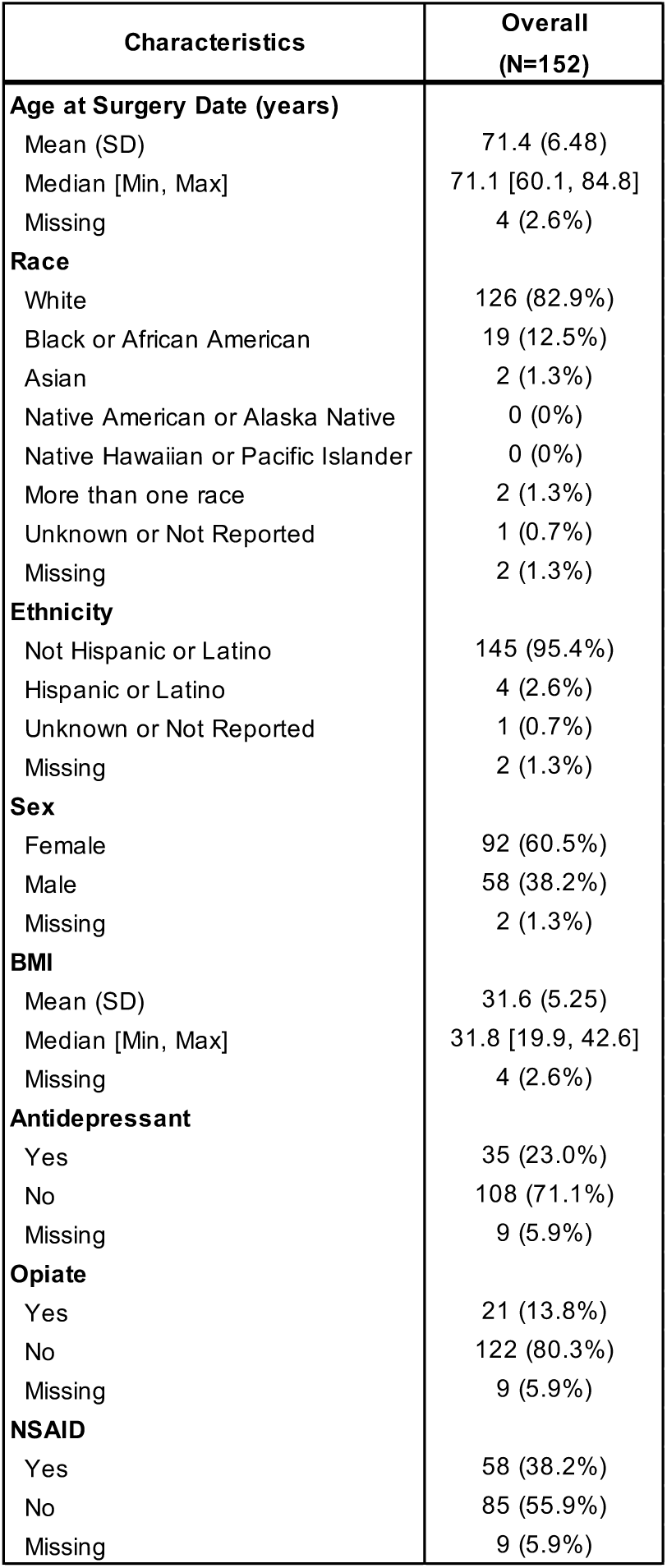
PRIME cohort characteristics.

Biomarker concentrations and assay performance in plasma and stimulated (TruCulture) samples are provided in **Table S1**. Intra-assay CVs (1.2-11.1%) and inter-assay CVs (range 0.9-38.4%) were generally excellent (<10%). LASSO stably selected biomarkers across all four resilience outcomes are shown in **Table S2**. The sample size varied by analysis based on who had available outcome data for the ERD / RT of interest (**Tables S2A** and **S2B**, respectively). In plasma, all but 5 of the biomarkers were selected in a stable set for at least one resilience outcome (**Table S3**). The most frequently selected plasma biomarker for ERD was CD30, and for RT was leptin (**Figure 1** and **Table S3**). A small subset of biomarkers demonstrated perfect stability scores in plasma (appearing in all 10 of 10 stable sets), but were never selected in the LPS- or FLU-stimulated models including: IFN-α2a, IL-12p70, IL-5, MIP-1β (pain intensity ERD); IL-5, IL-8 (pain interference ERD) and IL-5 (pain interference RT); IFN-γ, IL-1β, IL-2 (step count ERD) and fibrinogen, IFN-γ, IL-1β, IL-4 (step count RT); and IFN-γ, IL-13 (LE-PADL ERD) and IL-10 (LE-PADL RT). The plasma biomarkers most strongly positively and negatively associated with ERD and RT, considering the linear regression effect size after adjusting for all other biomarkers were: IFN-γ (positive association with function [LE-PADL] ERD), IL-5 (negative association with pain [pain interference] ERD), MCP-1 (positive association with function [LE-PADL] RT), and TNFR1 (negative association with function [LE-PADL] RT) (**Figure S1A** for ERD and **S1B** for RT).

**Figure 1.**
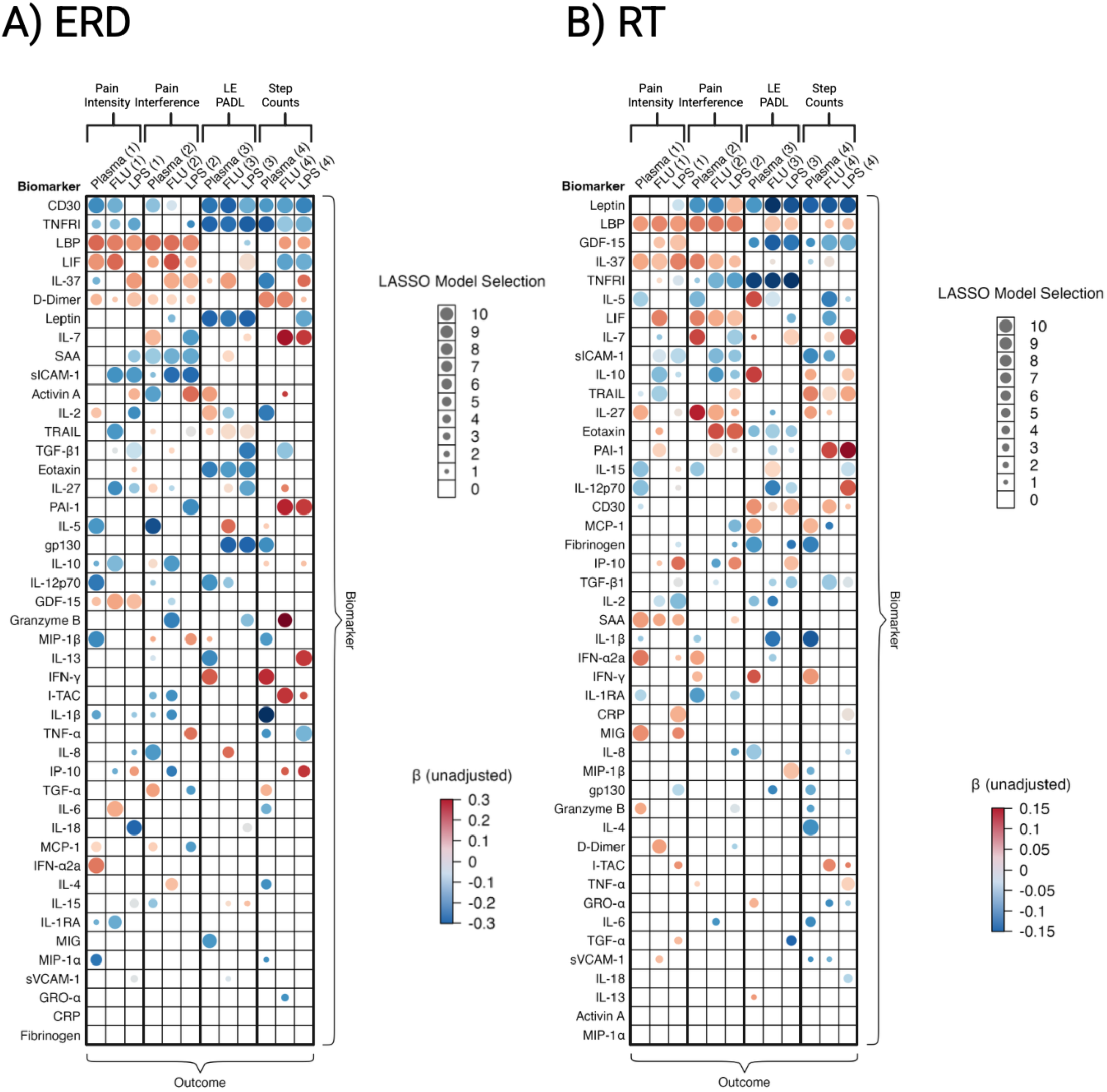
Bubble heatmaps of biomarker associations with resilience outcomes across Plasma, FLU, and LPS absolute contexts. Resilience outcomes, A) Expected Recovery Differential (ERD) and B) Resilience Trajectory (RT) are shown as single, aggregated bubble heatmaps including 12 biomarker–outcome contexts (3 biomarker parametrizations × 4 outcomes). Rows represent biomarkers, ordered by descending total stable set selections (sum of stability score across the 12 displayed contexts). Columns represent the biomarker parametrizations modeled for each resilience outcome and correspond to Plasma concentrations and, for the LPS and influenza stimulation paradigms, absolute 24 hr stimulated concentrations (LPS and FLU; for ratio and percent-change parametrizations see **Figure S3**). Four outcome types are depicted: (1) Pain Intensity, (2) Pain Interference, (3) LE PADLs, and (4) Step Counts. Within each biomarker-context cell, bubble size (legend: “LASSO Model Selection”) denotes the number of times that biomarker was selected by LASSO models within that specific context (0–10, where 10 indicates selection in the stable set for all 10 analytic configurations). Bubble color represents the unadjusted regression coefficients, with blue indicating negative associations and red indicating positive associations.

### Blood-based biomarkers induced by LPS and FLU antigens

Compared to NULL (sample incubated 24 hours at 37°C without immune stimulant), the majority of analytes were stimulated significantly by either FLU uniquely (eight analytes: PAI-1, TGF-β1, IL-2, Fibrinogen, LIF, sICAM-1, sVCAM-1 and gp130), LPS uniquely (eleven analytes: Granzyme B, IFN-γ, IL-1β, IL-6, IL-10, IL-12p70, IL-37, TNF-α, I-TAC, IL-27, TRAIL), or both (20 analytes: Activin A, IL-4, MIG, IFN-α2a, IL-1Ra, IL-5, IL-13, IL-15, IP-10, MIP-1α, MIP-1β, eotaxin, IL-7, TGF-α, GRO-α, IL-8, MCP-1, D-dimer, IL-18 and TNFRI) demonstrating that these two stimuli probe overlapping but distinct aspects of immune responsiveness (**Figure 2, Table S3** and **SHINY app** at https://primeknee.shinyapps.io/prime_viz/ representing interactive dilution-corrected cytokine plots).

**Figure 2.**
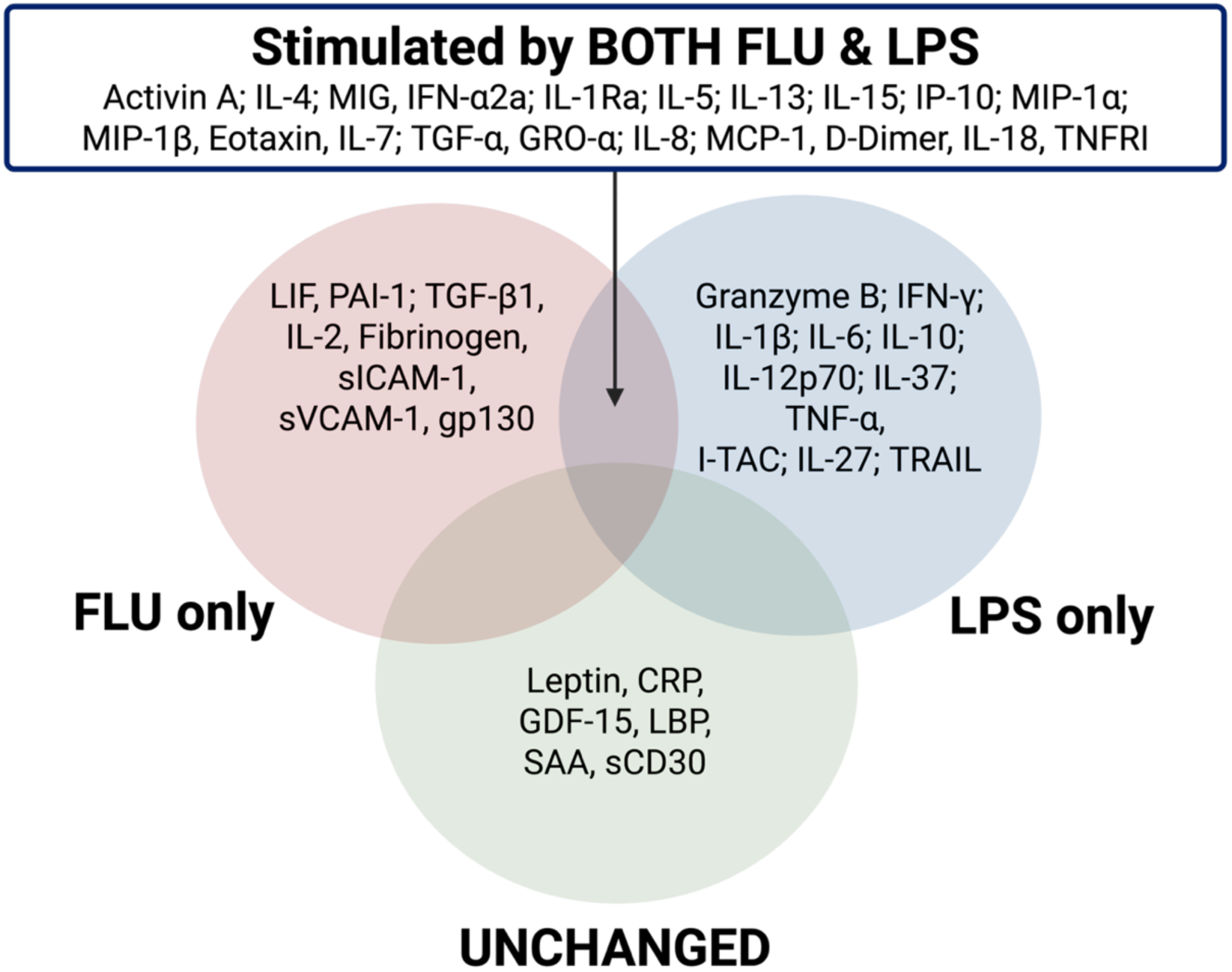
Patterns of biomarker stimulation *ex vivo*. Biomarkers were classified into four groups based on statistically significant changes relative to the NULL condition (24 h incubation at 37°C without stimulant): unchanged, stimulated by influenza antigen (FLU) only, lipopolysaccharide (LPS) only, or both FLU and LPS following 24 h *ex vivo* stimulation at 37°C. The majority of analytes were stimulated by at least one condition. Data visualization at https://primeknee.shinyapps.io/prime_viz/. **Biomarker Abbreviations**: Activin A (Activin A), CRP (C-reactive protein), D-Dimer (D-dimer), Eotaxin (Eotaxin/CCL11), Fibrinogen (Fibrinogen), GDF-15 (Growth/differentiation factor 15), GRO-α (Growth-regulated oncogene-alpha/CXCL1), Granzyme B (Granzyme B), gp130 (Glycoprotein 130/IL-6 receptor subunit beta), IFN-α2a (Interferon alpha-2a), IFN-γ (Interferon gamma), I-TAC (Interferon-inducible T-cell alpha chemoattractant/CXCL11), IL-1β (Interleukin-1 beta), IL-1Ra (Interleukin-1 receptor antagonist), IL-2 (Interleukin-2), IL- 4 (Interleukin-4), IL-5 (Interleukin-5), IL-6 (Interleukin-6), IL-7 (Interleukin-7), IL-8 (Interleukin-8/CXCL8), IL-10 (Interleukin-10), IL-12p70 (Interleukin-12 p70 heterodimer), IL-13 (Interleukin-13), IL-15 (Interleukin-15), IL-18 (Interleukin-18), IL-27 (Interleukin-27), IL-37 (Interleukin-37), IP-10 (Interferon gamma–induced protein 10/CXCL10), LBP (Lipopolysaccharide-binding protein), Leptin (Leptin), LIF (Leukemia inhibitory factor), MCP-1 (Monocyte chemoattractant protein-1/CCL2), MIG (Monokine induced by interferon-gamma/CXCL9), MIP-1α (Macrophage inflammatory protein-1 alpha/CCL3), MIP-1β (Macrophage inflammatory protein-1 beta/CCL4), PAI-1 (Plasminogen activator inhibitor-1), SAA (Serum amyloid A), sCD30 (Soluble CD30), sICAM-1 (Soluble intercellular adhesion molecule-1), sVCAM-1 (Soluble vascular cell adhesion molecule-1), TGF-α (Transforming growth factor-alpha), TGF-β1 (Transforming growth factor-beta 1), TNF-α (Tumor necrosis factor-alpha), TNFRI (Tumor necrosis factor receptor I), and TRAIL (TNF-related apoptosis-inducing ligand).

Importantly, the 24-hour NULL condition itself was not static. Several biomarkers increased over 24 hours at 37°C (CD30, I-TAC, IL-27, TRAIL, Eotaxin, IL-7, TGF-α, PAI-1, TGF-β1), while others decreased (CRP, GDF-15, LBP, SAA, D-dimer, IL-18, Fibrinogen, sICAM-1, sVCAM-1, TNFR1, gp130). These changes indicate that some analytes are actively secreted by circulating immune cells during incubation, whereas others may be consumed, processed, or degraded under *ex vivo* conditions. To account for this dynamic baseline, all stimulated values were compared directly to the 24-hour NULL condition, and an analyte was defined as “stimulated” only when it differed significantly from this time-matched control.

The specificity of this peripheral blood cell assay system was further supported by the absence of LPS-or FLU-induced stimulation of CRP, SAA, LBP and GDF-15 (all primarily liver-derived), leptin (adipose produced), or sCD30, which reflects T-cell activation *in vivo* and is unlikely to be released within the 24-hour incubation of this whole blood *ex vivo* assay. Overall, the majority of analytes demonstrated robust immune activation, expanding the biological dynamic range and enabling discrimination of stimulus-specific response patterns.

### A Core Set of Biomarkers Predicts Resilience

The top stable biomarker of each resilience outcome—ERD and RT for Pain Intensity, Pain Interference, LE PADLs, and Step Counts—were selected using the exploratory stability selection framework as described in Methods. Heatmaps (**Figure 1**) depict the stability score, or total number of times each biomarker was selected among the ‘top ten’ as well as the direction of association (red positive, blue negative) from the unadjusted linear regression models across the resilience measures. Of the 45 analytes tested in traditional plasma, FLU-stimulated (absolute values), or LPS-stimulated samples (absolute values), every analyte was selected at least once for at least one of the outcomes. Each biomarker had 240 possible selection opportunities into interim stable sets (10 selections across 4 domains, 2 resilience measures and three analyte types); five biomarkers—LBP, leptin, TNFR1, CD30, and LIF—were selected in ≥50% of these opportunities (**Table S3**). We observed substantial overlap between biomarkers selected for ERD and those selected for RT outcomes; LBP (in all samples) and IL-5 and IFN-γ (in plasma samples) were selected >50% of the time for both ERD and RT outcomes.

Some biomarkers (CRP, IL-18, IP-10, PAI-1 and TGF-β1) were only selected from LPS- or FLU-stimulated plasma; among these, IL-18 was exclusively selected from LPS-stimulated plasma, while no biomarkers were exclusively selected from FLU-stimulated plasma. A core subset of 11 biomarkers—CD30, D-dimer, eotaxin, GDF-15, IL-37, LBP, leptin, LIF, SAA, TNFRI and TRAIL—were repeatedly selected across multiple analytic conditions (**Figure 1**), including plasma, LPS-stimulated, and FLU-stimulated samples, suggesting stimulus-agnostic relevance to resilience. For ERD outcomes, the most selected analytes (summing across all analytic methods of modeling concentrations) were as follows: LBP for pain intensity; LBP and SAA for pain interference; CD30, Eotaxin, TNFR1 and Leptin for LE-PADLs; and TNFR1 for Step Counts. The most selected analytes for RT outcomes were as follows: IL-37 for pain intensity; LBP and Leptin for pain interference; TNFR1 for LE-PADLs; and Leptin and TNFR1 for Step Counts.

### Pain- vs. Function-Specific Biomarker Associations

Considering unstimulated plasma measures, 58% (26 of 45) of biomarkers were stably selected for both pain and function outcomes (for at least one parameterization and resilience type), while the remainder showed domain specificity; I-TAC, IFN-α2a, IL-15, IL-1Ra, LBP, LIF and SAA were selected exclusively for pain outcomes, whereas CRP, eotaxin, fibrinogen, gp130, GRO-α, IL-4, IL-6 and sVCAM-1 were specific to functional outcomes. These latter associations are consistent with the role of gp130 in IL-6 trans-signaling and tissue repair relevant to physical recovery(17). Across all biomarker measures (plasma, FLU- and LPS-stimulated), IL-1Ra consistently predicted ERD and RT pain outcomes, but not functional outcomes, while GRO-α was associated exclusively with function.

Together, these distinct biomarker sets define immune signatures differentiating pain versus functional resilience outcomes (**Figure 1** and **Graphical Abstract**).

### STRING Network and Functional Enrichment Analysis

The distinction between pain- and function-associated biomarkers was reinforced at the pathway level by STRING network analysis (**Figure S2**). Biomarkers selected across all ten LASSO iterations were organized into six networks derived by combining two outcome domains (pain versus function) with three stimulation conditions (plasma, LPS- and FLU-stimulated). All networks demonstrated strong protein-protein interaction (PPI) enrichment (p <1.0 × 10⁻¹⁶), indicating that selected biomarkers are more interconnected than expected by chance and share a common cytokine and immune driven framework (**Figure S2A**).

Despite this shared foundation, domain- and stimulus-specific KEGG enrichment patterns emerged (**Figure S2B**). Pain networks were characterized by enrichment of innate immune recognition and intracellular signaling pathways. Toll-like receptor signaling was enriched exclusively in the plasma pain network (FDR=2.4 × 10⁻¹⁰), while NF-kappa B signaling was enriched across all pain networks but absent from function networks. The plasma pain network was further uniquely enriched for RIG-I-like receptor and chemokine signaling pathways, consistent with an interferon/chemokine-centered signature.

In contrast, function networks were enriched for pathways linking immune signaling with coagulation and tissue repair. Complement and coagulation cascades (FDR=8.3 × 10^−5^) and platelet activation (FDR=0.007) were enriched exclusively in the FLU-stimulated function network. The function network from plasma samples showed strong enrichment for Th1/Th2 cell differentiation (FDR=4.8 × 10⁻¹¹) and IL-17 signaling (FDR=5.4 × 10⁻^9^), along with T cell receptor signaling and Fc epsilon RI signaling, reflecting greater engagement of adaptive immunity and thrombo-inflammatory processes in functional recovery.

Stimulus-specific distinctions further refined these patterns. The pain network from LPS stimulated samples was uniquely enriched for TGF-β and AGE-RAGE signaling, implicating profibrotic and metabolic stress pathways, whereas the pain network from FLU stimulated samples was uniquely enriched for apoptosis and natural killer cell-mediated cytotoxicity, consistent with cell death and innate lymphocyte activation. Collectively, these findings suggest that pain-related resilience signals are driven primarily by innate immune recognition and chemokine/interferon polarization, while function-related resilience additionally engages thrombo-inflammatory, coagulation, and adaptive immune pathways.

These distinctions are shaped by the type of immune stimulus applied. Notably, because the analyte panel was predefined based on known immune relevance, these STRING analyses are inherently constrained to this targeted set; unbiased proteomic approaches may reveal additional pathways and interactions. Overall, these results demonstrate that *ex vivo* immune responsiveness captures biologically meaningful and reproducible signals associated with surgical resilience, with distinct patterns emerging by stimulation condition, outcome domain, and biomarker parameterization.

### Direction of Biomarker Associations With Resilience Outcomes

Several of the most frequently selected biomarkers—such as LIF, IL-37, IL-7, TRAIL, and PAI-1—showed both positive and negative associations with resilience in unadjusted regression models depending on the specific outcome (**Figure 1**). This reversal was even more pronounced when comparing results across all analyte parameterization methods (**Figure S3**). In contrast, a small set of biomarkers showed consistent associations across outcomes and all parameterization approaches: LBP and IL-13 were reliably positively associated with resilience, whereas TGF-β1, TNFR1, fibrinogen, and sICAM-1 were reliably negatively associated. Adjusted effect estimates—derived from models including all other biomarkers in the outcome-specific stable selection set—and for RT outcomes, an indicator for intercurrent illness—did not meaningfully alter the direction or magnitude of associations of these particular biomarkers relative to unadjusted estimates (**Figure S1**).

### Ceiling Effects Underlie Reversals in Biomarker–Outcome Association Direction

For some biomarkers, the direction of association with outcomes depended on how stimulation responses were parameterized—whether as absolute concentrations, ratios to the null condition, or percent change. We compared six representations: absolute 24-hour LPS- or FLU-stimulated concentrations; stimulated-to-NULL ratios; and percent change relative to the NULL condition. To investigate reversals in association direction across parameterizations, we used Bland–Altman plots of stimulated–minus–NULL concentrations versus NULL concentrations (interactive Bland-Altman plots available at https://primeknee.shinyapps.io/prime_viz/). A directional reversal was defined as opposite associations observed when using absolute stimulated concentrations versus ratio and/or percent-change measures. A significant negative slope in these plots indicates a ceiling effect, in which higher baseline (null) concentrations are associated with smaller stimulated-minus-null differences. This compression can attenuate—or even reverse—associations when biomarkers are expressed as ratios or percent change rather than absolute stimulated concentrations. Across 90 Bland–Altman analyses (45 biomarkers × 2 stimulation conditions), reversals were observed for 11 biomarkers: LIF, IL-27, IL-37 (LPS and FLU); LBP, CRP, PAI-1 (LPS only); I-TAC, sCD30, IL-5, SAA, eotaxin (FLU only). Of these, 9 were consistent with a ceiling effect: LIF (LPS and FLU); LBP, CRP and PAI-1 (LPS); and I-TAC, sCD30, IL-27, IL-37 and SAA (FLU). The remaining two reversals lacked evidence of a ceiling effect.

### Explained Variance (R²) Attributable to Stable Biomarker Sets

Variance explained by stable biomarker sets was assessed using cross-validated R². Overall, biomarkers explained more variance in function than pain outcomes, consistently across ERD and RT metrics and for both the full stable sets (**Table 2A**) and those with perfect stability scores (10/10 selections) (**Table 2B**). The 29 baseline clinical parameters used to derive ERD (the ‘ERD derivation model’) accounted for 30–37% of pain variance and 38–60% of function variance in the raw outcomes (in-sample R²). These models were optimized for prediction, whereas stable biomarker sets were designed to identify reproducible signals associated with resilience.

**Table 2.**
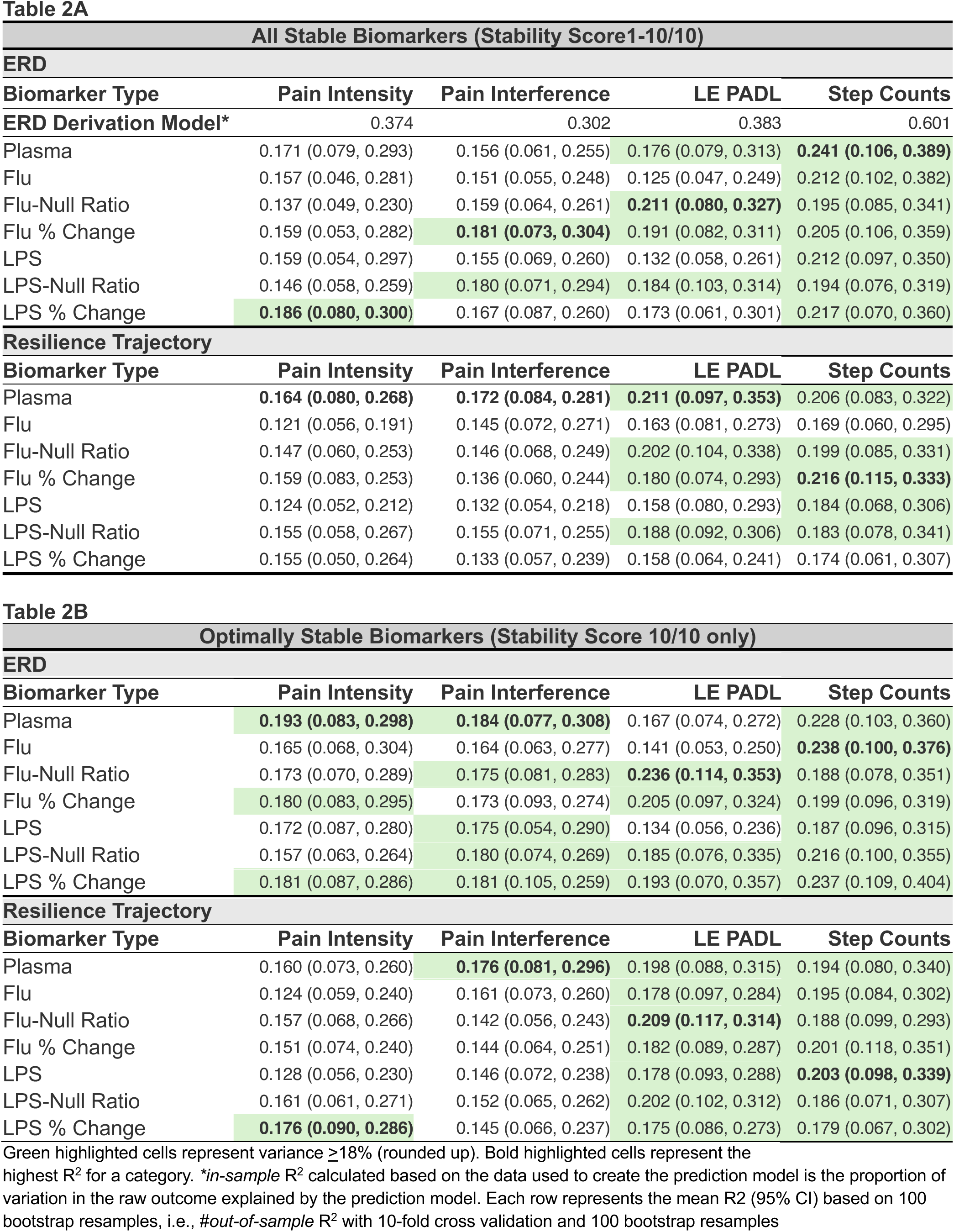
Cross-validated R² comparisons for ERD and RT outcomes.

Despite this, baseline biomarkers explained additional variance in resilience outcomes, accounting for 14–19% of residual variance in pain ERD and 13–24% in function ERD outcomes (**Table 2A**). For RT outcomes—defined by postoperative trajectories rather than clinical residuals—biomarkers explained 12–17% of pain variance and 16–22% of function variance (**Table 2A**). Plasma values alone consistently explained ≥16% of variance across ERD and RT outcomes and yielded the highest R^2^ for half of all outcomes (ERD and RT for pain intensity, pain interference, LE PADL and step counts) (**Table 2A**). However, comparisons of plasma R² to other analyte parameterizations showed no significant differences (95% empirical CIs), indicating similar performance across approaches. Notably, the more parsimonious subset of perfectly stable biomarkers performed comparably or better than the full stable set in 63% of all analyses (**Table 2B versus 2A**).

## Discussion

Understanding why some older adults recover rapidly after surgery while others experience persistent pain or functional decline is central to precision perioperative medicine. In this study of older adults undergoing TKA, we evaluated circulating and *ex vivo*–stimulated immune biomarkers as predictors of eight resilience-related pain and functional outcomes.

Using a standardized whole blood *ex vivo* stimulation system, three principal findings emerged. First, a stable reproducible subset of biomarkers—including CD30, LBP, leptin, LIF, TNFRI, SAA, GDF-15, D-dimer, eotaxin, IL-37, and TRAIL—was consistently associated with resilience across stimulation conditions and outcomes. Several cytokines (IL-4, IL-6, IL-7, IL-13, IL-15) showed uniformly positive associations with recovery, whereas others (IL-1β, IL-5, IFN-α2a, GDF-15) were predominantly negative. Second, LPS and FLU stimulation revealed distinct immune activation signatures, indicating that the choice of stimulant is not interchangeable. Third, the magnitude—and in some cases the direction—of biomarker–outcome associations depended strongly on how stimulated responses were parameterized. Directional reversals occurred for 11 biomarkers and 9 of these were consistent with a ceiling effect, underscoring that analytic representation can materially influence biological interpretation.

We observed robust and stimulus-specific cytokine induction. LPS responses were consistent with activation of innate immune signaling via pattern-recognition receptors such as Toll-like receptor 4, whereas FLU stimulation reflected antigen-specific adaptive immune responsiveness. The stimulus-specificity of several biomarkers suggests that these assays capture complementary dimensions of immune capacity relevant to surgical recovery.

Whole-blood stimulation assays offer practical advantages, including preservation of physiological cell–cell interactions and minimal preprocessing. However, they primarily capture early immune responses and may be influenced by recent infections, vaccination, or short-term immune fluctuations.

Unexpectedly, FLU stimulation induced lower Granzyme B responses than LPS. One potential explanation is that 85% of participants had received influenza vaccination within the prior year, potentially placing them near maximal FLU-specific responsiveness at baseline. However, Bland–Altman analysis did not support a clear ceiling effect for this cytokine. Notably, the plasma and stimulated biomarkers examined here performed similarly in several contexts, suggesting that these static and dynamic immune measures generally reflect similar underlying biology.

Biomarkers explained meaningful variance in resilience beyond clinical predictors. Clinical characteristics accounted for 30–60% of variance in raw recovery (ERD), with biomarkers explaining an additional 13–24% of residual variance from that explained by the clinical (ERD) prediction model. For RT outcomes, biomarkers alone explained 12–21% of variance. Stimulated biomarkers—particularly FLU-based measures—tended to better predict functional recovery, whereas plasma biomarkers performed comparably for pain outcomes. This divergence was mirrored in pathway analyses: pain-associated biomarkers were enriched for innate immune and chemokine/interferon signaling, whereas function-associated biomarkers were enriched for coagulation, platelet activation, and adaptive immune pathways. These findings suggest that pain and functional resilience reflect partially distinct biological processes.

Strengths of this study include a well-characterized surgical cohort, comprehensive biomarker profiling under multiple stimulation conditions, and rigorous analytic approaches, including correction of technical variation in TC sample dilution using a urea-based method. The use of two distinct immune challenges enabled interrogation of complementary aspects of immune function. Limitations include lack of normalization to immune cell counts, and reliance on a targeted biomarker panel. We did not apply the LASSO-based stability selection framework across combined biomarker sets (plasma, FLU-, and LPS-stimulated) to prioritize clinical feasibility, as requiring multiple specimen types would limit scalability. However, integrating these datasets may have improved predictive performance. External validation will be essential to confirm generalizability.

## Conclusions

Circulating and *ex vivo*–stimulated immune biomarkers capture biologically meaningful, outcome-specific variation in postoperative resilience among older adults undergoing TKA. Dynamic immune responses to LPS and FLU provide complementary mechanistic insight and, in some cases, incremental predictive value—particularly for functional recovery—whereas traditional plasma biomarkers perform comparably for several outcomes with greater clinical feasibility. The distinct immune signatures underlying pain versus functional recovery further indicate that resilience reflects partially separable biological processes. Validation in independent cohorts is needed, but these results support the potential for both static and dynamic immune measures to inform precision perioperative strategies in aging populations.

## Methods

### PRIME Cohort

PRIME is a prospective cohort study of community-dwelling adults aged ≥60 years who underwent elective unilateral TKA at Duke University, All study procedures were approved by the Duke University Institutional Review Board, and the study was conducted in accordance with the principles of the Declaration of Helsinki. All participants provided written informed consent (13). The trial was registered as NCT04235309. Eligibility required English proficiency and access to a Bluetooth-enabled mobile device for actigraphy. Exclusion criteria included inability to ambulate independently (with or without an assistive device), known dementia or failed telephone cognitive screen, receipt of medical treatment for non-skin cancer within the prior 12 months, and severe vision or hearing impairments that precluded reliable cognitive assessment or telephone interviews despite accommodation. Within 6 weeks prior to TKA surgery, participants completed standard preoperative assessments (self-reported demographics, body mass index, comorbidities and medications from the electronic health record) along with preoperative measures of physical, psychological, social, and cognitive reserve.

Physical reserve measures included a 3-minute walk test, and grip strength using a hand dynamometer. Psychological reserve measures included the PHQ-9 and Brief Resilience Scale. Social determinants and resource-based measures of reserve included education level, the PROMIS Emotional Support scale, and a self-reported measure of financial resource sufficiency. Cognitive reserve measures included the 3MS, Trails A and B tests, and the 15-item recall test. Outcome data were collected at baseline, as well as several timepoints post-surgery: daily in the week post-surgery, and at the 1-, 2-, 4-, and 6-month post-surgical timepoints. The outcomes included self-reported measures of pain intensity and pain interference (PROMIS), lower extremity activities of daily living (LE PADL), and average daily step counts derived from Garmin-watch collected data. Data were also collected at the 1-, 2-, 4-, and 6-month timepoints on post-surgery intercurrent events including falls, COVID-19, and participant report of an illness or injury that prevented regular activities for at least one day during the recovery period.

### Definitions of resilience measures

PRIME considered two complementary constructs of resilience—ERD and RT—as previously described (14, 15, 18). For both measures and across all outcomes, we coded higher values to indicate better recovery. ERD quantifies a participant’s observed postoperative recovery at a specific timepoint relative to their individualized predicted recovery. ERD represents the individual-level residual—the portion of postoperative outcome variance not explained by 29 clinical characteristics (**Table S4**), including demographics, baseline reserve measures, surgical factors, and baseline status for the corresponding domain (e.g., baseline Pain Intensity for the Pain Intensity ERD). Negative residuals indicate worse-than-predicted recovery, whereas positive residuals indicate better-than-predicted recovery. ERD was calculated at 6 months for pain intensity and pain interference, at 4 months for step counts, and at 1 month for LE-PADLs. These timepoints were selected to maximize inter-individual variability in resilience—capturing recovery before outcomes plateaued—while minimizing missing data. For example, step count ERD was calculated at 4 months due to peak Garmin watch participation, whereas LE-PADL ERD was calculated at 1 month because scores reached a ceiling thereafter.

In contrast, RT characterizes overall recovery through 6 months by integrating longitudinal assessments at 1, 2, 4, and 6 months into latent trajectory classes identified using latent class trajectory analysis. Unlike the ERD approach, RT analysis takes into account measures from all available time points for each individual. Continuous RT outcomes were derived by summing the posterior probabilities of membership in “more resilient” trajectory classes for each outcome; RT was defined as the posterior probability of classification into a highly resilient recovery trajectory class.

Specifically, resilient classes were defined as follows: for pain intensity, Class 1 (High Resilience); for pain interference, Classes 1 or 2 (High and Moderate Resilience); for LE PADLs, Classes 1 or 2 (Never Disabled and Fast Complete Recovery); and for step counts, Classes 3 or 4 (Modest and High Activity Gains).

### TruCulture blood collection system

We collected EDTA plasma, and 1 ml whole blood in 3 ml TruCulture® (TC) tubes containing 2 ml RPMI 1640 (liquid cell culture medium) with NaCitrate anticoagulant (proprietary TruCulture media, Rules Based Medicine (RBM), Austin, TX) either without stimulants (RBM #782-001086) for the NULL condition, or with lipopolysaccharide (LPS 0.01 ng/ml, RBM # 782-001087), or high dose influenza antigen (Sanofi Fluzone 5ug/ml, custom produced by RBM). TC samples were incubated for 24 hours at 37°C, gently ‘plunged’ with Seraplas filters (Sarstedt Cat No 53.677) to separate supernatant from cells, and cell-free supernatants were stored at −80C until analysis. Long-term stability of the custom-manufactured Fluzone TruCulture® tubes was confirmed by the manufacturer (RBM) using an IL-2 endpoint. For this analysis, Fluzone TruCulture samples from two healthy donors were stored at −20°C for 0 (3-5 days), or 3-, 6-, 12- and 18-months. IL-2 concentrations of stored samples were compared to those frozen a total of only 3-5 days. Frozen samples yielded stable IL-2 concentrations over the 18-month interval with ratios of stored samples to baseline (stored 3-5 days) ranging from 1.01 - 1.40. Traditional EDTA plasma samples were centrifuged (3000 rpm, 15 minutes, 4°C) and stored at −80°C until analysis.

### Dilution factor determinations based on urea concentrations

Urea concentrations of plasma and TC samples were measured using the UniCel DxC 600/800 System and Synchron Systems Multi Calibrator, an automated Beckman clinical chemistry analyzer (Beckman Instruments, Brea, CA) with change in absorbance at 340 nm, allowing for the detection of urea. A Human Control Plasma (HCP) was measured multiple times throughout the run, demonstrating excellent low intra-assay variability (3.6%).

Dilution factors (DFs) were calculated as the ratio of plasma urea/TC urea of matched samples. To assess the ability of urea concentration to correct for experimentally induced dilution, six unique human plasma samples were prepared at no dilution (neat), 2-, 3-, 4-, and 5-fold dilutions in RPMI media, thereby simulating the experimental dilution conditions of the TC samples. Although the actual (pre-determined) DFs of these six samples were significantly positively correlated with the experimentally derived DFs based on measured urea concentrations (R^2^ 0.99, p<0.0001), we noticed that the experimentally derived DFs underestimated the actual DFs, particularly at the higher DFs (**Figure S4**). Therefore, this bias was corrected based on the linear fit of actual to predicted DFs:

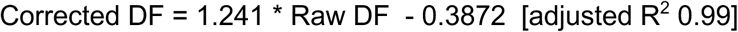

A total of 12 samples had no matched plasma sample. The TruCulture sample urea concentrations corresponding to these were imputed using the overall mean dilution factor of 2.89; this was a reasonable assumption given the consistency of urea concentrations of the measured samples and the few outliers in the sample set as a whole.

### Biomarker measurement by ELISA

A total of 45 blood-based biomarkers, 33 pro-inflammatory and 12 anti-inflammatory, were measured in matched plasma and TruCulture samples (**Graphical Abstract** and **Figure 1**). Custom multiplex assays were designed to optimize sample use: MSD UPLEX Custom Proinflammatory 10-plex (IFN-γ, IL-1β, IL-2, IL-4, IL-5, IL-6, IL-8, IL-10, IL-12p70, and IL-13; 2-fold dilution), MSD UPLEX Custom Cytokine 10-plex (Eotaxin, IFNα2a, IL-7, IL-15, IL-18, IP-10, MCP-1, MIP-1α, MIP-1β, TNF-α; 2-fold dilution), MSD UPLEX/RPLEX Custom 10-plex (sCD30, Granzyme B, GRO-α, IL-1Ra, IL-27, IL-37, I-TAC, Leptin, MIG, TRAIL; 2-fold dilution), MSD UPLEX Custom 2-plex (gp130, TNFRI; 100-fold dilution), MSD RPLEX Custom 2-plex (LIF, TGF-α; undiluted), MSD VPLEX Vascular Injury (Cat#K1511198D: CRP, ICAM-1, SAA, VCAM-1; 1000-fold dilution), MSD RPLEX GDF-15 (Cat#K151YDR; 100-fold dilution), MSD RPLEX LBP (Cat#K151K5R; 500-fold dilution), MSD UPLEX TGF-β (Cat#K151XWK; 1.34-fold dilution), D-dimer (Biomedica Diagnostics, Cat#602; 21-fold dilution), Fibrinogen (Hyphen Biomed, Cat#RK024A; 200,000-fold dilution), PAI-1 (Bio-Techne, Cat#DSE100; 2-fold dilution), Activin A (Bio-Techne, Cat#DAC00B; 5-fold dilution). The Molecular Measures Core (MMC) performed all immunoassays within the Duke Biomarkers Core Facility, housed in the Duke Molecular Physiology Institute.

### Biomarker selection and statistical modeling

The primary goal of the statistical analysis was to identify preoperative biomarkers most robustly associated with resilience outcomes. Both resilience measures—the ERD and the RT—were evaluated across four domains: pain intensity, pain interference, LE PADLs, and step counts. Biomarkers were analyzed across 7 derived types, including plasma values; LPS- and FLU-stimulated concentrations; their NULL-normalized ratios and percent changes. All values derived from TruCulture were dilution-corrected. Values below the lower limit of detection (LLOD) were imputed as one-half the LLOD. Biomarkers were log-transformed and standardized, except percent-change measures, which were cube-root transformed.

Predictive biomarkers were identified using an exploratory stability-selection framework using LASSO, with 200 resamples for each of 10 analytic configurations (19). For each resample, LASSO was applied using one of five penalty parameters (targeting ∼5–25 non-zero coefficients) under two resampling strategies—bootstrap (20) and complementary pairs (21)—yielding 10 combinations of penalty parameter and resampling strategy per analysis. Biomarkers within a configuration were ranked by the number of resamples in which they were selected, and the top 10 formed the configuration-specific interim stable set. Biomarkers were then given a stability score, representing the number of interim stable sets in which they appeared, and those selected at least once were carried forward into a ‘grand stable set’. For each selected biomarker in the grand stable set, we fit unadjusted linear regression models and adjusted models controlling for all selected biomarkers and an indicator for intercurrent illness to assess effect direction. Statistical uncertainty was not reported due to the data-driven selection process. Predictive performance was evaluated using out-of-sample R² from 10-fold cross-validation repeated with 100 bootstrap resamples. The mean and 95% empirical confidence interval for the out-of-fold R² were computed for each biomarker type, outcome, and resilience measure. All analyses were conducted in R version 4.4.0 (22).

## Acknowledgments

We wish to thank Dr Michael Muehlbauer for urea analyses and Mr Nikolay D Perumov for assistance with ELISA analyses.

## Author Contributions

Formal Analysis: MA, SP

Funding Acquisition: HEW, CSC-E, VBK, SP

Investigation: JLH

Methodology: VBK, NDG, MA, JLH, AB, SP, CS, HEW, CSC-E

Project Administration: VBK, SP, HEW, CSC-E

Resources: HEW, CSC-E, MA, SP

Software: MA, SP

Supervision: VBK, SP, HEW, CSC-E

Validation: MA, JLH

Visualization: NDG, AB, MA

Writing: VBK, NDG

Writing – Review & Editing: VBK, NDG, MA, JLH, AB, SP, CS, HEW, CSC-E

## Data Availability

Data from this study are not publicly available, but de-identified datasets can be requested via email to the PRIME principal investigators (HEW, CSC-E).

## Related manuscripts

None

## Supporting Information Captions

**Figure S1. Forest plots of top biomarkers**. Beta estimates across biomarker types and by resilience outcome (ERD vs RT). Plots show unadjusted and adjusted effects for biomarkers in the grand stable sets for plasma, LPS, and FLU models. Adjusted estimates were derived from regression models that included all other biomarkers from that outcome’s stable set of top selections and an indicator for intercurrent illness. Panels are organized by resilience outcome (pain, interference, LE PADL, step counts).

**Figure S2. STRING protein–protein interaction networks and KEGG pathway enrichment for LASSO-selected biomarkers predicting surgical resilience outcomes. (A)** STRING networks constructed from biomarkers selected 10/10 times by LASSO in both ERD and RT models. <u>Left column</u>: Pain Outcomes (Pain Intensity + Pain Interference combined). <u>Right column</u>: Function Outcomes (LE PADLs + Step Counts combined). <u>Rows</u>: Plasma (top), LPS-stimulated absolute concentrations (middle), FLU-stimulated absolute concentrations (bottom). Nodes represent proteins; edges represent known and predicted protein–protein interactions at medium confidence (interaction score ≥ 0.400). All networks showed PPI enrichment p < 1.0 × 10⁻¹⁶. **(B)** KEGG pathway enrichment across the six networks. Each cell shows the number of input proteins found in that pathway (observed/background); color intensity reflects FDR significance (see legend in the graphic). Dashes indicate the pathway was not enriched (FDR >0.05) in that network. Pathways are grouped by biological theme. “Str.” (Best Strength) = highest log₁₀(observed/expected) across networks; “Best FDR” = lowest false discovery rate across all six networks; “#” (Nets Enriched) = number of networks in which the pathway reached FDR < 0.05. Infectious disease pathways that are enriched primarily because they contain cytokine/chemokine genes in their KEGG definitions are omitted for clarity.

**Figure S3. Comprehensive bubble heatmap summary of biomarker associations with resilience outcomes across all biomarker parametrizations.** Resilience outcomes, **A)** Expected Recovery Differential (ERD) and **B)** Resilience Trajectory (RT) are shown as single, aggregated bubble heatmaps. <u>Rows</u> represent biomarkers, ordered by descending sum of LASSO model selections across all biomarker parametrizations and outcomes. <u>Columns</u> represent the various biomarker parametrizations modeled for each resilience outcome. Column names correspond to the seven biomarker parametrizations evaluated for each biomarker: plasma concentrations (Plasma); for LPS and influenza stimulation paradigms, A = absolute 24 hr stimulated concentrations (LPS-A, FLU-A), B = stimulated-to-null ratio at 24 hr (LPS-B, FLU-B), and C = percent change at 24 hr (LPS-C, FLU-C). Four outcome types are depicted: (1) Pain Intensity, (2) Pain Interference, (3) LE PADLs, and (4) Step Counts. Within each biomarker-context cell, bubble size denotes the LASSO selection frequency, and bubble color denotes the unadjusted regression coefficient (β), with blue indicating negative associations and red indicating positive associations (scale shown).

**Figure S4. Correlation between actual and experimentally derived dilution factors based on urea concentrations.** Measurements from the clinical chemistry analyzer slightly overestimated urea concentrations at sample dilutions greater than twofold leading to modest underestimation of the experimentally derived dilution factors. To address this bias, corrected dilution factors were calculated using the best-fit line (equation shown in the figure) relating actual to experimentally derived dilution factors. The overall mean dilution factor was 2.89-fold, slightly below the expected 3-fold dilution; this value was used to impute corrected analyte concentrations for the twelve individuals for whom dilution factors could not be calculated due to missing baseline plasma samples. Graphic created in BioRender.

**Table S1. Biomarker concentrations and assay performance in plasma and TruCulture samples.** The four numeric columns (Plasma, 24-hr Null, 24-hr FLU, 24-hr LPS) report mean concentration with SD in parentheses. 24-hr Null, 24-hr FLU, and 24-hr LPS were corrected for dilution using the urea method. For each biomarker, we ran a repeated-measures ANOVA (within-subject factor = sample type); Tukey’s HSD post-hoc tests were performed only if the omnibus ANOVA reached α = 0.05. The significance symbols in the “Significant 24h inducer” column show which pairwise comparisons were significant and the direction of change: A = 24-Hour LPS-Stim vs. 24-Hour Null; B = 24-Hour Flu-Stim vs. 24-Hour Null; C = 24-Hour LPS-Stim vs. 24-Hour Flu-Stim; D= 24-Hour Null vs. Plasma. A plus sign **(+)** after the letter indicates the first condition listed in the comparison was significantly higher than the second; a minus sign (**−**) indicates the first condition was significantly lower. The numeric value in the parentheses is the Tukey HSD p-value (two-sided). *None* indicates that ANOVA gave p <0.05 but no pairwise comparisons were significant after Tukey correction; *NS* indicates that ANOVA gave p >0.05. Intra-/inter-assay %CVs are reported as “intra/inter”. All reported TruCulture concentration (NULL, LPS-and FLU-stimulated) are corrected for dilution based on the urea method as described in the Methods. For comprehensive visualization, all six possible pairwise comparisons are displayed graphically in the interactive Shiny application (https://primeknee.shinyapps.io/prime_viz/), which presents the full set of comparisons for each biomarker alongside the underlying distributions. Activin A (Activin A), CRP (C-reactive protein), D-Dimer (D-dimer), Eotaxin (Eotaxin/CCL11), Fibrinogen (Fibrinogen), GDF-15 (Growth/differentiation factor 15), GRO-α (Growth-regulated oncogene-alpha/CXCL1), Granzyme B (Granzyme B), gp130 (Glycoprotein 130/IL-6 receptor subunit beta), IFN-α2a (Interferon alpha-2a), IFN-γ (Interferon gamma), I-TAC (Interferon-inducible T-cell alpha chemoattractant/CXCL11), IL-1β (Interleukin-1 beta), IL-1Ra (Interleukin-1 receptor antagonist), IL-2 (Interleukin-2), IL-4 (Interleukin-4), IL-5 (Interleukin-5), IL-6 (Interleukin-6), IL-7 (Interleukin-7), IL-8 (Interleukin-8/CXCL8), IL-10 (Interleukin-10), IL-12p70 (Interleukin-12 p70 heterodimer), IL-13 (Interleukin-13), IL-15 (Interleukin-15), IL-18 (Interleukin-18), IL-27 (Interleukin-27), IL-37 (Interleukin-37), IP-10 (Interferon gamma–induced protein 10/CXCL10), LBP (Lipopolysaccharide-binding protein), Leptin (Leptin), LIF (Leukemia inhibitory factor), MCP-1 (Monocyte chemoattractant protein-1/CCL2), MIG (Monokine induced by interferon-gamma/CXCL9), MIP-1α (Macrophage inflammatory protein-1 alpha/CCL3), MIP-1β (Macrophage inflammatory protein-1 beta/CCL4), PAI-1 (Plasminogen activator inhibitor-1), SAA (Serum amyloid A), sCD30 (Soluble CD30), sICAM-1 (Soluble intercellular adhesion molecule-1), sVCAM-1 (Soluble vascular cell adhesion molecule-1), TGF-α (Transforming growth factor-alpha), TGF-β1 (Transforming growth factor-beta 1), TNF-α (Tumor necrosis factor-alpha), TNFRI (Tumor necrosis factor receptor I), and TRAIL (TNF-related apoptosis-inducing ligand).

**Table S2**. **Table S2A. Stably-selected PBMC-stimulated biomarkers associated with ERD measures of resilience after knee surgery. Table S2B.** Stably-selected PBMC-stimulated biomarkers associated with RT measures of resilience after knee surgery. The table lists all biomarkers selected by LASSO for the in at least one of ten separate analytic configurations (using all combinations of 2 sampling methods and 5 penalty parameters termed the Grand Stable Set) as predictors of resilience outcomes (**S2A**) ERD and (**S2B**) RT. Biomarkers in **bold** font were selected all 10 times/iterations. LASSO selection was performed on biomarkers from traditional plasma and stimulated whole blood whose values were defined in one of three manners: 1) absolute biomarker concentrations of the stimulated samples; 2) stim/null ratios, defined as the ratio of the 24-hour stimulated value to its paired 24-hour unstimulated control; and 3) percent change, defined as (24-hour stimulated minus 24-hour unstimulated)/24-hour unstimulated. Prior to LASSO selection, all stimulated biomarker concentrations were corrected for the dilution factors inherent in the TruCulture tube collection system, using the urea method. Color coding is based on unadjusted beta estimates of biomarkers and outcomes: **red** positive association, **black** negative association; n represents number of individuals in the specific analysis.

**Table S3. Frequency and percentage of biomarker selection by resilience outcome.** Outcomes are ERD, RT, ERD+RT across ALL measures (LPS, FLU and plasma in bold **black** font) and for plasma (PL in **red** font) alone. Ranked by overall frequency of selection. ERD and RT equating to 240 possible chances of selection overall (*ALL from plasma, LPS or FLU stimulated) or 80 possible chances of selection in plasma (PL). ERD=Expected Recovery Differential; RT=Resilience Trajectory

**Table S4. The twenty-nine clinical predictors used to construct the expected recovery differential (ERD) outcome.** All measures derived from baseline visit except for age derived at the time of surgery.

## References

1. Jayabalan P, Gustafson J, Sowa GA, Piva SR, Farrokhi S. A Stimulus-Response Framework to Investigate the Influence of Continuous Versus Interval Walking Exercise on Select Serum Biomarkers in Knee Osteoarthritis. Am J Phys Med Rehabil. 2019;98(4):287–91.

2. Abadir PM, Bandeen-Roche K, Bergeman C, Bennett D, Davis D, Kind A, et al. An overview of the resilience world: Proceedings of the American Geriatrics Society and National Institute on Aging State of Resilience Science Conference. Journal of the American Geriatrics Society. 2023;71(8):2381–92.

3. Walston J, Varadhan R, Xue QL, Buta B, Sieber F, Oni J, et al. A Study of Physical Resilience and Aging (SPRING): Conceptual framework, rationale, and study design. Journal of the American Geriatrics Society. 2023;71(8):2393–405.

4. Müller S, Kröger C, Schultze JL, Aschenbrenner AC. Whole blood stimulation as a tool for studying the human immune system. Eur J Immunol. 2024;54(2):e2350519.

5. Riyazi N, Slagboom E, de Craen AJ, Meulenbelt I, Houwing-Duistermaat JJ, Kroon HM, et al. Association of the risk of osteoarthritis with high innate production of interleukin-1beta and low innate production of interleukin-10 ex vivo, upon lipopolysaccharide stimulation. Arthritis and rheumatism. 2005;52(5):1443–50.

6. Goekoop RJ, Kloppenburg M, Kroon HM, Frolich M, Huizinga TW, Westendorp RG, et al. Low innate production of interleukin-1beta and interleukin-6 is associated with the absence of osteoarthritis in old age. Osteoarthritis and cartilage. 2010;18(7):942–7.

7. McElhaney JE, Verschoor CP, Andrew MK, Haynes L, Kuchel GA, Pawelec G. The immune response to influenza in older humans: beyond immune senescence. Immun Ageing. 2020;17:10.

8. Jorda A, Eberl S, Nussbaumer-Pröll A, Sarhan M, Weber M, Tegrovsky LE, et al. Reproducibility of LPS-Induced ex vivo Cytokine Response of Healthy Volunteers Using a Whole Blood Assay. J Inflamm Res. 2024;17:4781–90.

9. Hall MW, Knatz NL, Vetterly C, Tomarello S, Wewers MD, Volk HD, et al. Immunoparalysis and nosocomial infection in children with multiple organ dysfunction syndrome. Intensive Care Med. 2011;37(3):525–32.

10. Svanberg R, MacPherson C, Zucco A, Agius R, Faitova T, Andersen MA, et al. Early stimulated immune responses predict clinical disease severity in hospitalized COVID-19 patients. Commun Med (Lond). 2022;2:114.

11. Nikkhoo B, Mohammadi M, Hasani S, Sigari N, Borhani A, Ramezani C, et al. Elevated interleukin (IL)-6 as a predictor of disease severity among Covid-19 patients: a prospective cohort study. BMC Infect Dis. 2023;23(1):311.

12. McElhaney JE, Verschoor CP, Haynes L, Pawelec G, Loeb M, Andrew MK, et al. Key Determinants of Cell-Mediated Immune Responses: A Randomized Trial of High Dose Vs. Standard Dose Split-Virus Influenza Vaccine in Older Adults. Front Aging. 2021;2.

13. Whitson HE, Crabtree D, Pieper CF, Ha C, Au S, Berger M, et al. A template for physical resilience research in older adults: Methods of the PRIME-KNEE study. J Am Geriatr Soc. 2021;69(11):3232–41.

14. Colón-Emeric C, Pieper CF, Schmader KE, Sloane R, Bloom A, McClain M, et al. Two Approaches to classifying and quantifying physical resilience in longitudinal data. J Gerontol A Biol Sci Med Sci. 2020;75(4):731–8.

15. Colón-Emeric CS, Peskoe S, Ashner MC, Kraus VB, Huebner JL, Hall KS, et al. Clinical Predictors of Resilience following Total Knee Arthroplasty: the PRIME-KNEE Study. J Gerontol A Biol Sci Med Sci. 2026;81(5):glag086.

16. Kraus VB, Huebner JL, Fink C, King JB, Brown S, Vail TP, et al. Urea as a passive transport marker for arthritis biomarker studies. Arthritis Rheum. 2002;46(2):420–7.

17. Rose-John S. IL-6 trans-signaling via the soluble IL-6 receptor: importance for the pro-inflammatory activities of IL-6. Int J Biol Sci. 2012;8(9):1237–47.

18. Whitson H, Ashner M, Kraus V, Peskoe S, Huebner J, Simon C, et al. Comparing two methods to quantify physical resilience. J Frailty & Aging. 2026;in press.

19. Tibshirani R. Regression Shrinkage and Selection Via the Lasso. Journal of the Royal Statistical Society: Series B (Methodological). 1996;58:267–88.

20. Efron B, Tibshirani R. An Introduction to the Bootstrap. New York: Chapman and Hall/CRC; 1994.

21. Shah R, Samworth R. Variable Selection with Error Control: Another Look at Stability Selection. Journal of the Royal Statistical Society Series B: Statistical Methodology. 2013;75(1):55–80.

22. Team RC. The R Project for Statistical Computing2024. Available from: https://www.r-project.org/.

