## Supplementary Figures and Tables for "Immune Biomarker Signatures as Predictors of Functional and Pain Recovery After Total Knee Arthroplasty in Older Adults"

### PRIME PBMC Supplementary Figures
#### Figure S1

##### A) Forest Plot of biomarker beta estimates for (ERD)

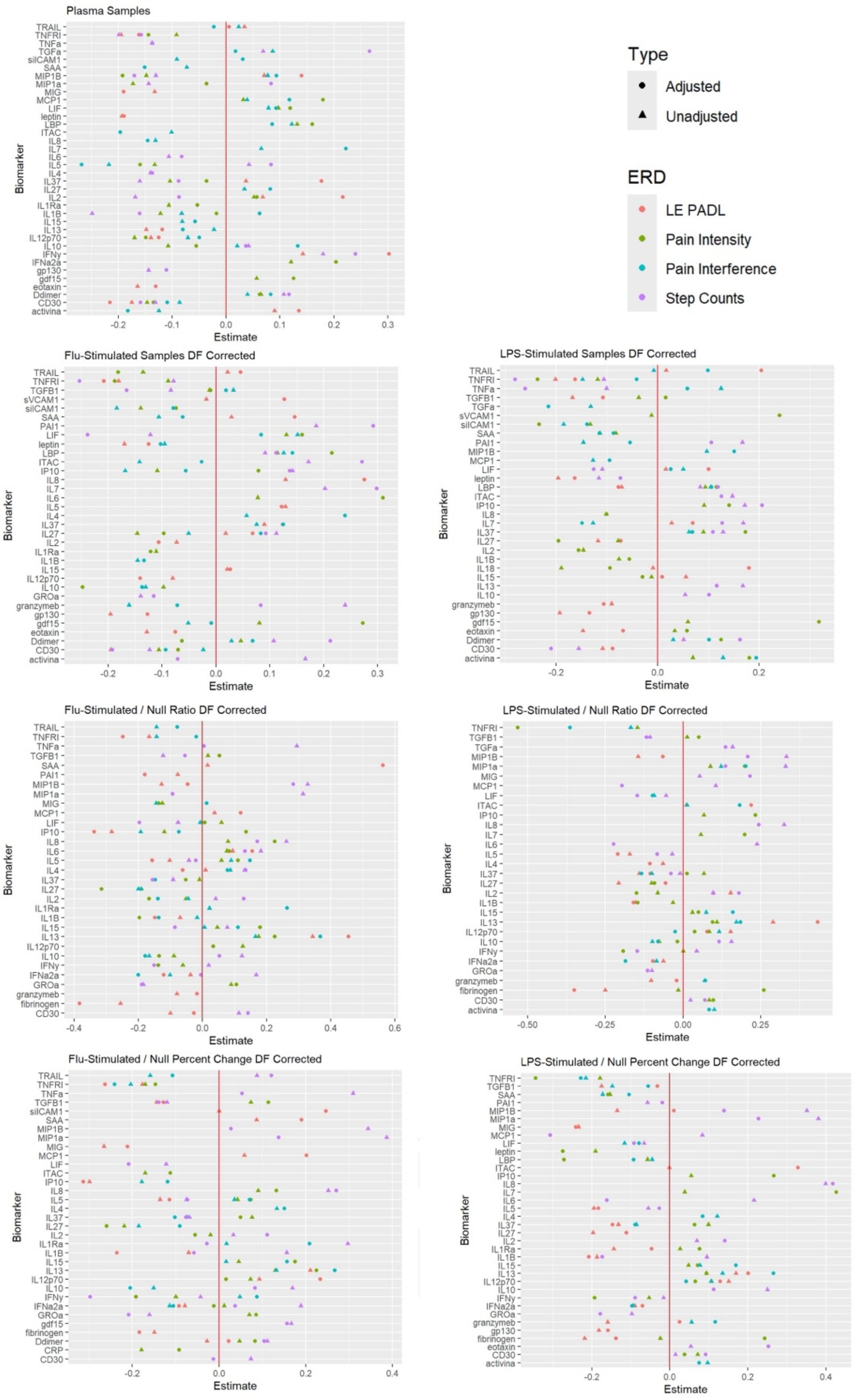

#### B) Forest Plot of biomarker beta estimates for (RT)

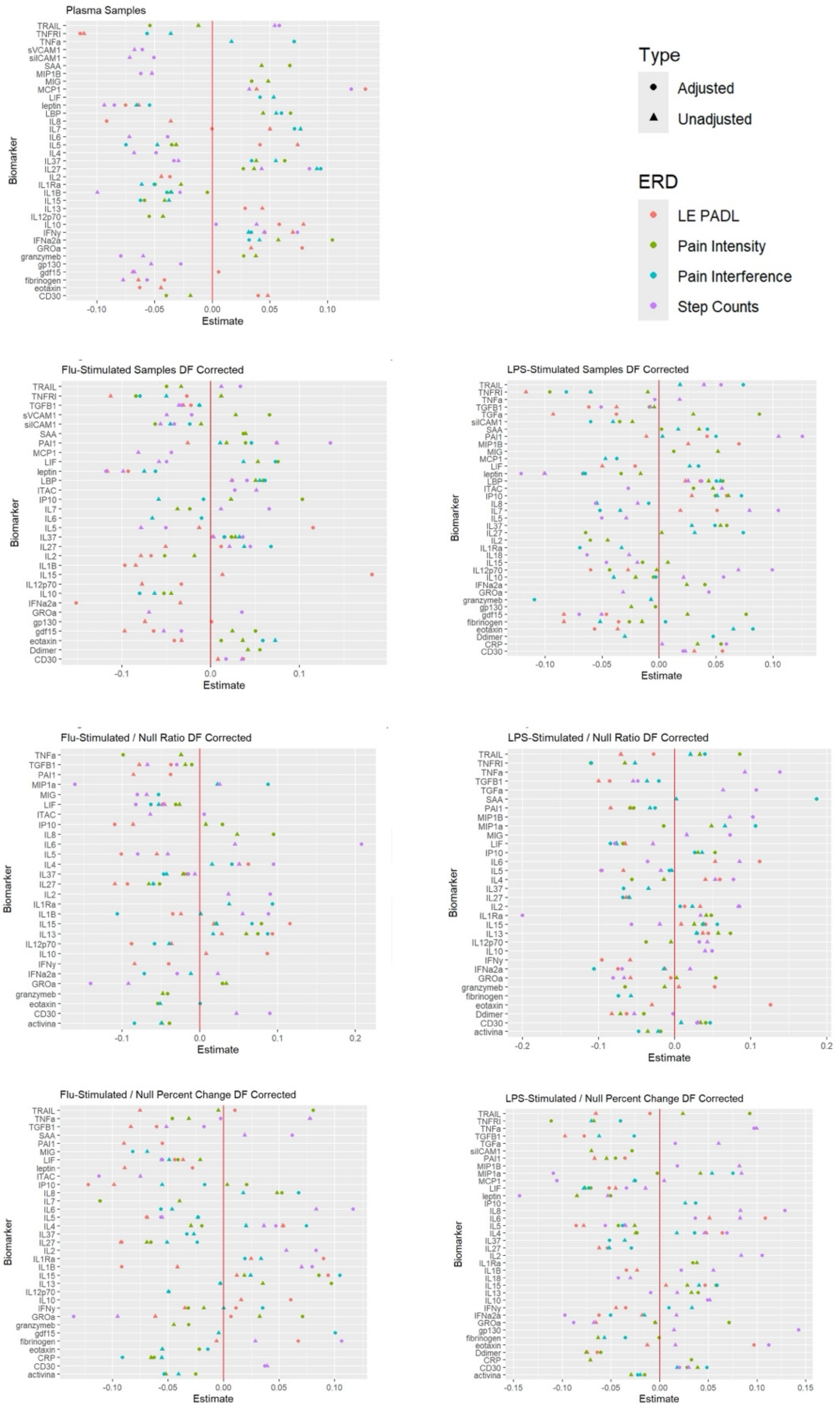

**Figure S1. Forest plots of top biomarkers**. Beta estimates across biomarker types and by resilience outcome, A) Expected Recovery Differential (ERD) and B) Resilience Trajectory (RT). Plots show unadjusted and adjusted effects for biomarkers in the grand stable sets for plasma, LPS, and FLU models. Adjusted estimates were derived from regression models that included all other biomarkers from that outcome's stable set of top selections and an indicator for intercurrent illness. Panels are organized by resilience outcome (pain, interference, lower extremity physical activities of daily living [LE PADL], step counts).

#### Figure S2

A

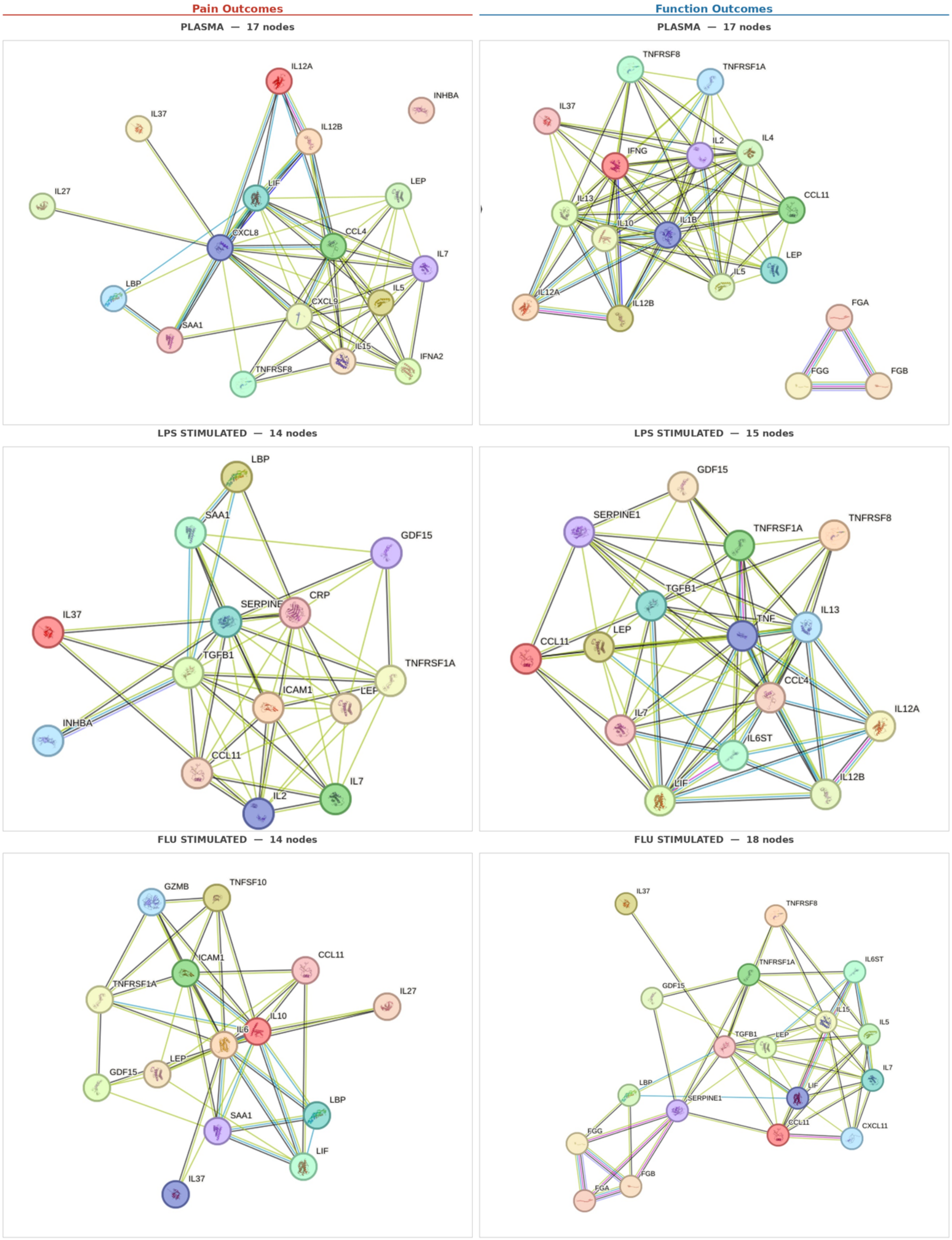

All networks: PPI enrichment $p < 1\times10^{-16}$ | Medium confidence (score ≥ 0.400)

**B**

#### KEGG Pathway Enrichment Across Six STRING Networks

**FDR:** ■$<10^{-15}$ ■$<10^{-10}$ ■$<10^{-5}$ ■$<10^{-3}$ ■<.01 □<.05 □n.s.

| KEGG Pathway | Pain Plas. | Pain LPS | Pain FLU | Func. Plas. | Func. LPS | Func. FLU | Str. | Best FDR | # |
|---|---|---|---|---|---|---|---|---|---|
| ***Core Cytokine & Immune Signaling*** | | | | | | | | | |
| Cytokine–cytokine receptor interaction | 15/282 | 9/282 | 10/282 | 14/282 | 14/282 | 13/282 | 1.81 | $1.5\times10^{-23}$ | 6/6 |
| JAK-STAT signaling | 8/158 | 3/158 | 4/158 | 9/158 | 7/158 | 6/158 | 1.82 | $3.5\times10^{-13}$ | 6/6 |
| Toll-like receptor signaling | 7/100 | – | – | – | – | – | 1.91 | $2.4\times10^{-10}$ | 1/6 |
| Viral protein interaction w/ cytokine & receptor | 4/96 | 4/96 | 6/96 | 5/96 | 5/96 | 5/96 | 1.94 | $8.1\times10^{-9}$ | 6/6 |
| C-type lectin receptor signaling | 2/101 | – | – | 5/101 | – | – | 1.76 | $5.5\times10^{-7}$ | 2/6 |
| RIG-I-like receptor signaling | 4/69 | – | – | – | – | – | 1.83 | $2.7\times10^{-5}$ | 1/6 |
| TNF signaling | 2/111 | – | 4/111 | – | – | 3/111 | 1.71 | $1.2\times10^{-4}$ | 3/6 |
| NF-κB signaling | 3/101 | 3/101 | 3/101 | – | – | – | 1.62 | 0.002 | 3/6 |
| Chemokine signaling | 3/186 | – | – | – | – | – | 1.27 | 0.009 | 1/6 |
| ***T/NK Cell & Adaptive Immunity*** | | | | | | | | | |
| Th1 & Th2 differentiation | 3/85 | – | – | 7/85 | – | – | 1.98 | $4.8\times10^{-11}$ | 2/6 |
| IL-17 signaling | 2/91 | – | – | 6/91 | – | – | 1.88 | $5.4\times10^{-9}$ | 2/6 |
| T cell receptor signaling | – | – | – | 5/100 | – | – | 1.76 | $5.5\times10^{-7}$ | 1/6 |
| Intestinal immune (IgA) | 2/43 | 2/43 | 2/43 | 4/43 | – | 3/43 | 2.03 | $1.2\times10^{-6}$ | 5/6 |
| Th17 differentiation | 2/91 | 2/99 | – | 4/99 | – | – | 1.67 | $2.2\times10^{-5}$ | 3/6 |
| Fc epsilon RI signaling | – | – | – | 3/65 | – | – | 1.73 | $3.1\times10^{-4}$ | 1/6 |
| NK cell cytotoxicity | – | – | 3/120 | – | – | – | 1.55 | 0.003 | 1/6 |
| Hematopoietic cell lineage | 2/90 | – | – | – | – | – | 1.41 | 0.034 | 1/6 |
| ***TGF-β, Coagulation & Metabolic*** | | | | | | | | | |
| Type I diabetes mellitus | 2/38 | – | – | 5/38 | 3/38 | – | 2.18 | $7.2\times10^{-9}$ | 3/6 |
| Complement & coagulation cascades | – | – | – | – | – | 4/82 | 1.73 | $8.3\times10^{-5}$ | 1/6 |
| FoxO signaling | – | – | 3/126 | – | – | – | 1.53 | 0.003 | 1/6 |
| Apoptosis | – | – | 3/131 | – | – | – | 1.51 | 0.003 | 1/6 |
| AGE-RAGE signaling | – | 3/96 | – | – | – | – | 1.64 | 0.003 | 1/6 |
| NAFLD | – | 3/146 | 3/146 | – | – | 3/146 | 1.46 | 0.004 | 3/6 |
| Platelet activation | – | – | – | – | – | 3/122 | 1.43 | 0.007 | 1/6 |
| Adipocytokine signaling | – | 2/68 | – | – | – | – | 1.62 | 0.028 | 1/6 |
| TGF-β signaling | – | 2/91 | – | – | – | – | 1.49 | 0.043 | 1/6 |
| ***Autoimmune & Inflammatory Disease*** | | | | | | | | | |
| Inflammatory bowel disease | 3/59 | 2/59 | – | 9/59 | 5/59 | – | 2.25 | $1.6\times10^{-16}$ | 4/6 |
| Allograft rejection | 3/34 | – | 2/34 | 7/34 | 3/34 | – | 2.38 | $2.1\times10^{-13}$ | 4/6 |
| Asthma | – | – | 2/27 | 5/27 | 3/27 | 2/27 | 2.33 | $2.1\times10^{-9}$ | 4/6 |
| Autoimmune thyroid disease | 2/48 | – | 2/48 | 4/48 | – | – | 1.98 | $1.7\times10^{-6}$ | 3/6 |
| Graft-versus-host disease | – | – | 2/36 | 3/36 | – | – | 1.98 | $6.4\times10^{-5}$ | 2/6 |
| Rheumatoid arthritis | 2/83 | 2/83 | – | – | – | – | 1.53 | 0.030 | 2/6 |

**Figure S2. STRING protein–protein interaction networks and KEGG pathway enrichment for LASSO-selected biomarkers predicting surgical resilience outcomes. (A)** STRING networks constructed from biomarkers selected 10/10 times by LASSO in both ERD and RT models. Left column: Pain Outcomes (Pain Intensity + Pain Interference combined). Right column: Function Outcomes (LE PADLs + Step Counts combined). Rows: Plasma (top), LPS-stimulated absolute concentrations (middle), FLU-stimulated absolute concentrations (bottom). Nodes represent proteins; edges represent known and predicted protein–protein interactions at medium confidence (interaction score ≥ 0.400). All networks showed PPI enrichment $p < 1.0 \times 10^{-16}$. **(B)** KEGG pathway enrichment across the six networks. Each cell shows the number of input proteins found in that pathway (observed/background); color intensity reflects FDR significance (see legend). Dashes indicate the pathway was not enriched (FDR >0.05) in that network. Pathways are grouped by biological theme. "Str." (Best Strength) = highest $\log_{10}$(observed/expected) across networks; "Best FDR" = lowest false discovery rate across all six networks; "#" (Nets Enriched) = number of networks in which the pathway reached FDR <0.05. Infectious disease pathways that are enriched primarily because they contain cytokine/chemokine genes in their KEGG definitions are omitted for clarity.

**Figure S3**

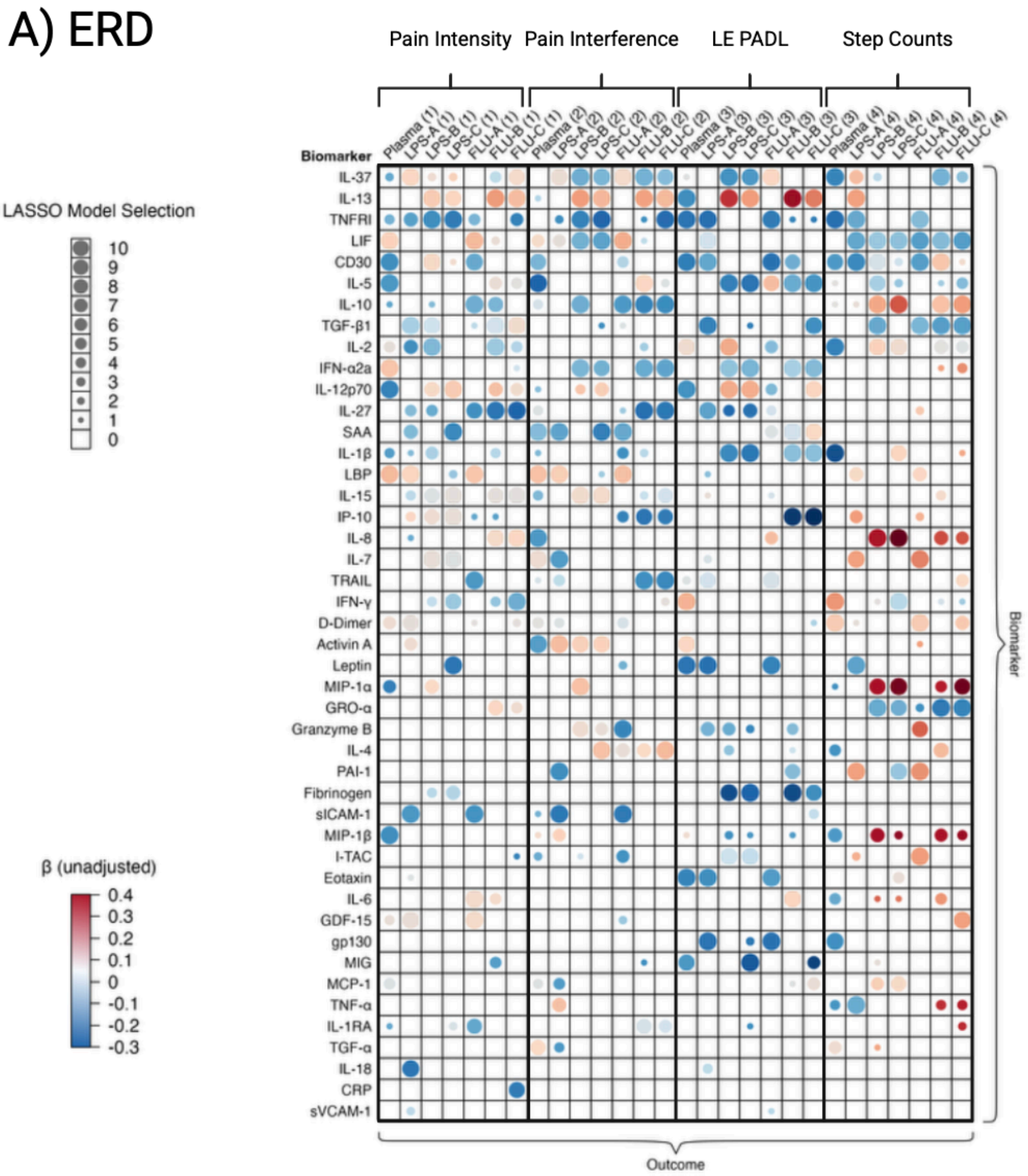

**Context codes**:
Plasma = plasma concentration
LPS-A/FLU-A = absolute 24-hr stimulated
LPS-B/FLU-B = stimulated-to-null ratio
LPS-C/FLU-C = percent change

**Outcome codes**:
(1) = Pain Intensity
(2) = Pain Interference
(3) = LE PADL
(4) = Step Counts

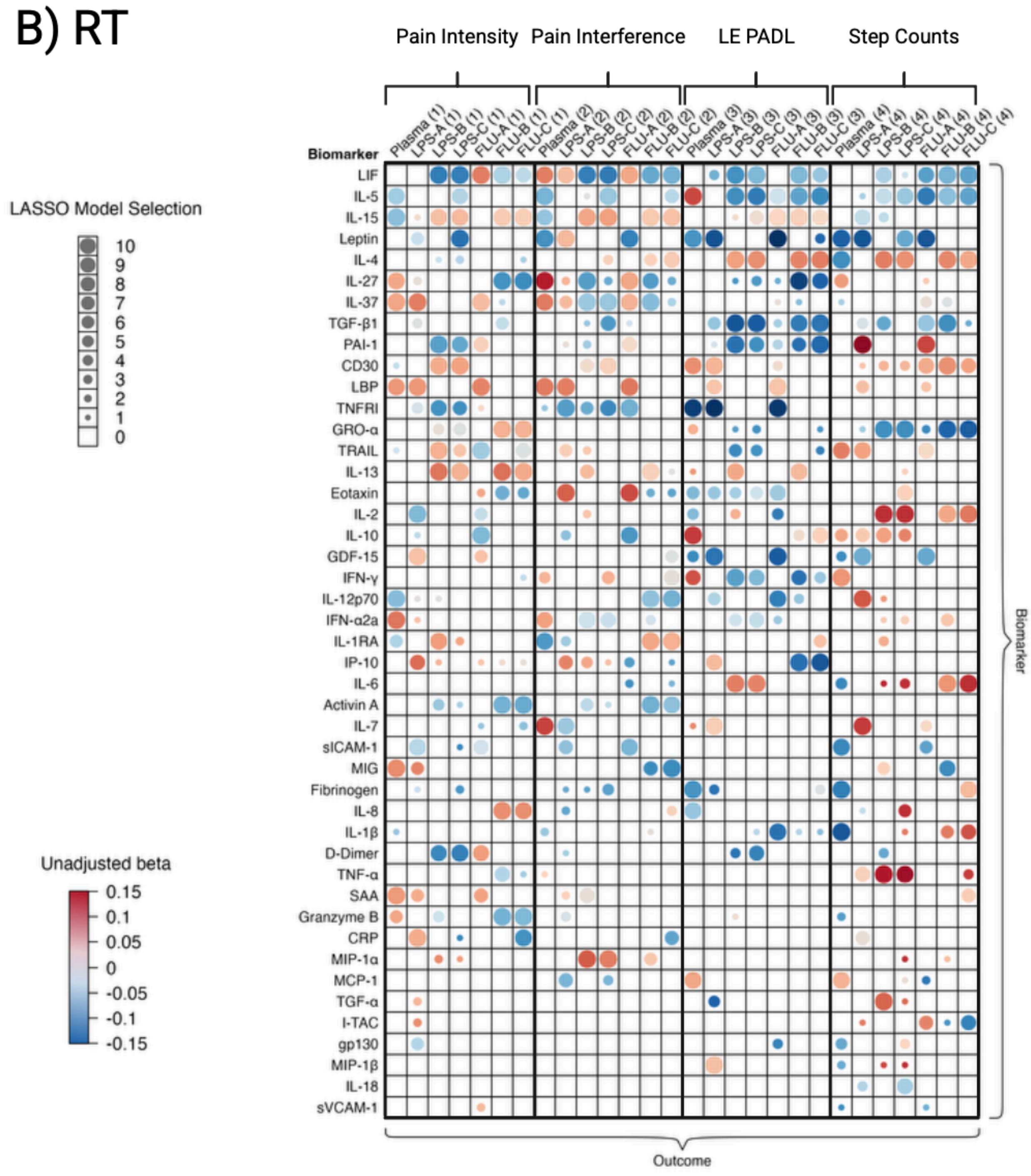

**Context codes**:
Plasma = plasma concentration
LPS-A/FLU-A = absolute 24-hr stimulated
LPS-B/FLU-B = stimulated-to-null ratio
LPS-C/FLU-C = percent change

**Outcome codes**:
(1) = Pain Intensity
(2) = Pain Interference
(3) = LE PADL
(4) = Step Counts

**Figure S3. Comprehensive bubble heatmap summary of biomarker associations with resilience outcomes across all biomarker parametrizations.** Resilience outcomes, panel **A**)

Expected Recovery Differential (ERD) and panel **B**) Resilience Trajectory (RT) are shown as single, aggregated bubble heatmaps. Rows represent biomarkers, ordered by descending sum of LASSO model selections across all biomarker parametrizations and outcomes. Columns represent the various biomarker parametrizations modeled for each resilience outcome. Column names correspond to the seven biomarker parametrizations evaluated for each biomarker: plasma concentrations (Plasma); LPS-A and FLU-A represent absolute 24 hr stimulated concentrations; LPS-B and FLU-B represent stimulated-to-null ratios at 24 hr; and LPS-C and FLU-C represent percent change at 24 hr. Four outcome types are depicted: (1) Pain Intensity, (2) Pain Interference, (3) LE PADLs, and (4) Step Counts. Within each biomarker-context cell, bubble size denotes the LASSO selection frequency, and bubble color denotes the unadjusted regression coefficient (β), with blue indicating negative associations and red indicating positive associations (scale shown).

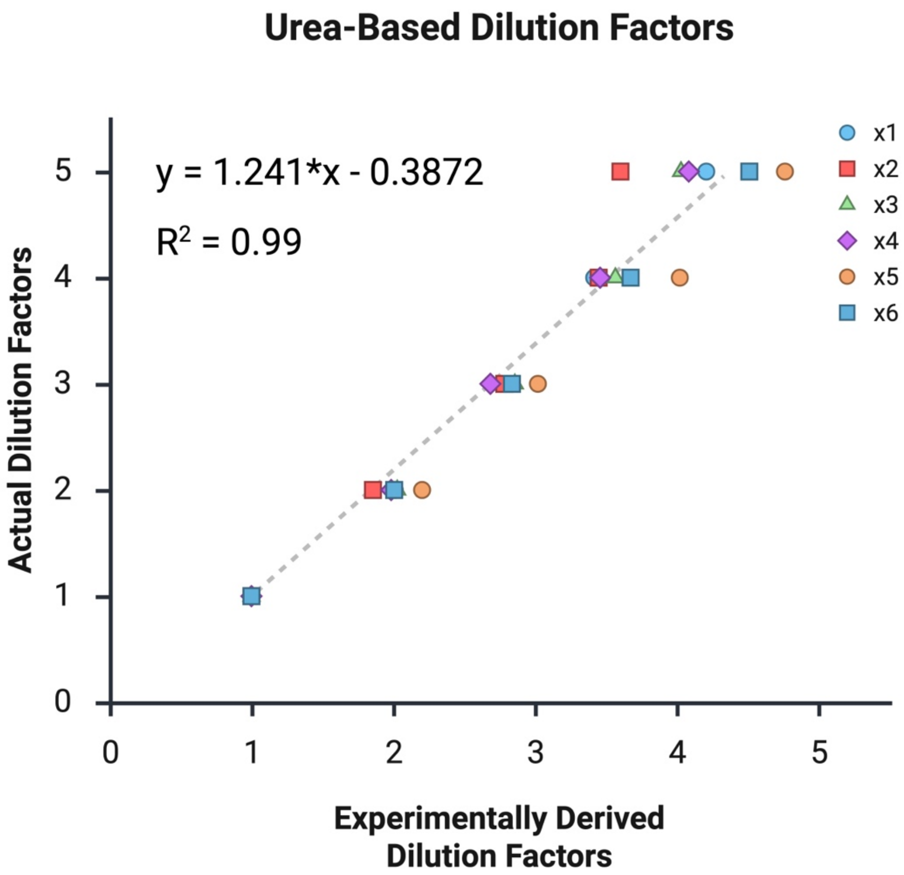

**Figure S4. Correlation between actual and experimentally derived dilution factors based on urea concentrations.** Measurements from the clinical chemistry analyzer slightly overestimated urea concentrations at sample dilutions greater than twofold leading to modest underestimation of the experimentally derived dilution factors. To address this bias, corrected dilution factors were calculated using the best-fit line (equation shown in the figure) relating actual to experimentally derived dilution factors. The overall mean dilution factor was 2.89-fold, slightly below the expected 3-fold dilution; this value was used to impute corrected analyte concentrations for the twelve individuals for whom dilution factors could not be calculated due to missing baseline plasma samples. Graphic created in BioRender.

**PRIME PBMC Supplementary Tables**

**Table S1. Biomarker concentrations and assay performance in plasma and TruCulture samples.**

| Biomarker | Biomarker Concentrations — mean (SD) | | | | Significant 24h inducer *(p value) | Intra-/inter-assay %CV |
|---|---|---|---|---|---|---|
| | Plasma | 24hr Null | 24hr LPS stimulated | 24hr FLU stimulated | | |
| Activin A | 291.99 pg/mL (131.68) | 195.04 pg/mL (196.0) | 2,002.23 pg/mL (976.36) | 1,781.01 pg/mL (1,540.66) | $A^+$ (<0.001) $B^+$ (<0.001) | 3.9 / 6.2 |
| CRP | 3,891.01 ng/mL (6,401.75) | 2,216.74 ng/mL (3676.92) | 2,206.73 ng/mL (3,605.99) | 2,244.41 ng/mL (3,625.92) | $D^-$ (<0.001) | 2.1 / 6.7 |
| D-Dimer | 597.18 ng/mL (342.60) | 344.89 ng/mL (310.52) | 454.88 ng/mL (490.04) | 489.75 ng/mL (429.69) | $A^+$ (<0.001) $B^+$ (<0.001) $D^-$ (<0.001) | 3.7 / 6.9 |
| Eotaxin | 292.07 pg/mL (126.48) | 2,394.67 pg/mL (1,215.64) | 2,579.05 pg/mL (1,206.55) | 2,735.63 pg/mL (1,352.00) | $A^+$ (0.046) $B^+$ (<0.001) $D^+$ (<0.001) | 8.7 / 3.7 |
| Fibrinogen | 4,012.25 µg/mL (958.98) | 2,175.20 µg/mL (727.19) | 2,243.73 µg/mL (638.74) | 2,390.79 µg/mL (955.54) | $B^+$ (0.002) $D^-$ (<0.001) | 3.7 / 7.2 |
| GDF-15 | 1,208.66 pg/mL (557.25) | 710.29 pg/mL (377.07) | 699.84 pg/mL (364.60) | 746.88 pg/mL (398.20) | $D^-$ (<0.001) | 3.7 / 5.2 |
| GRO-α | 107.59 pg/mL (115.24) | 1,140.56 pg/mL (541.58) | 8,836.16 pg/mL (4,372.47) | 14,428.66 pg/mL (16,505.93) | $A^+$ (<0.001) $B^+$ (<0.001) $C^-$ (<0.001) | 8.0 / 5.7 |
| Granzyme B | 2.99 pg/mL (2.22) | 51.33 pg/mL (119.49) | 4,728.40 pg/mL (3,512.51) | 191.89 pg/mL (318.42) | $A^+$ (<0.0001) $C^+$ (<0.0001) | 2.6 / 8.3 |
| gp130 | 262,120.28 pg/mL (46,075.76) | 160,259.51 pg/mL (40,511.35) | 163,169.13 pg/mL (34,359.32) | 178,664.83 pg/mL (37,817.69) | $B^+$ (<0.001) $C^-$ (<0.001) $D^-$ (<0.001) | 1.6 / 4.7 |
| IFN-α2a | 1.98 pg/mL (4.02) | 7.65 pg/mL (18.65) | 376.71 pg/mL (86.78) | 310.37 pg/mL (90.38) | $A^+$ (<0.001) $B^+$ (<0.001) $C^+$ (<0.001) | 8.7 / 3.5 |
| IFN-γ | 10.90 pg/mL (28.16) | 27.64 pg/mL (48.02) | 77,896.19 pg/mL (76,457.09) | 5,075.32 pg/mL (5,856.80) | $A^+$ (<0.001) $C^+$ (<0.001) | 1.9 / 10.1 |
| I-TAC | 18.85 pg/mL (18.26) | 793.7 pg/mL (1,969.60) | 1,438.25 pg/mL (2,787.37) | 926.23 pg/mL (1,671.13) | $A^+$ (<0.001) $C^+$ (<0.001) $D^+$ (<0.001) | 3.3 / 11.7 |
| IL-1β | 0.16 pg/mL (0.60) | 1.90 pg/mL (3.52) | 7,159.00 pg/mL (4,298.94) | 108.79 pg/mL (398.77) | $A^+$ (<0.001) $C^+$ (<0.001) | 4.2 / 3.1 |

| | | | | | | |
|---|---|---|---|---|---|---|
| IL-1Ra | 195.73 pg/mL (123.51) | 1,059.70 pg/mL (1,709.59) | 41,401.23 pg/mL (22,614.47) | 33,827.25 pg/mL (22,339.90) | $A^+$ (<0.001) $B^+$ (<0.001) $C^+$ (<0.001) | 10.1 / 4.8 |
| IL-2 | 0.48 pg/mL (0.54) | 1.04 pg/mL (1.59) | 91.28 pg/mL (60.75) | 632.31 pg/mL (712.65) | $B^+$ (<0.001) $C^-$ (<0.001) | 10.5 / 7.5 |
| IL-4 | 0.02 pg/mL (0.04) | 0.11 pg/mL (0.56) | 1.83 pg/mL (1.58) | 1.78 pg/mL (0.97) | $A^+$ (<0.001) $B^+$ (<0.001) | 7.5 / 0.9 |
| IL-5 | 0.91 pg/mL (3.00) | 0.80 pg/mL (2.80) | 14.07 pg/mL (5.10) | 12.89 pg/mL (5.50) | $A^+$ (<0.001) $B^+$ (<0.001) $C^+$ (0.006) | 2.5 / 38.4 |
| IL-6 | 1.77 pg/mL (1.46) | 3.45 pg/mL (6.12) | 64,722.13 pg/mL (25,921.88) | 570.87 pg/mL (3,660.92) | $A^+$ (<0.001) $C^+$ (<0.001) | 3.4 / 22.9 |
| IL-7 | 1.83 pg/mL (1.42) | 8.63 pg/mL (3.71) | 127.56 pg/mL (31.35) | 62.60 pg/mL (27.16) | $A^+$ (<0.001) $B^+$ (<0.001) $C^+$ (<0.001) $D^+$ (0.013) | 6.2 / 6.9 |
| IL-8 | 8.57 pg/mL (5.52) | 353.63 pg/mL (813.86) | 87,357.96 pg/mL (45,148.24) | 117,486.21 pg/mL (93,847.22) | $A^+$ (<0.001) $B^+$ (<0.001) $C^-$ (<0.001) | 2.9 / 10.3 |
| IL-10 | 0.99 pg/mL (7.50) | 0.58 pg/mL (3.68) | 338.91 pg/mL (252.04) | 12.31 pg/mL (23.76) | $A^+$ (<0.001) $C^+$ (<0.001) | 2.8 / 19.3 |
| IL-12p70 | 0.37 pg/mL (0.85) | 0.41 pg/mL (0.82) | 106.67 pg/mL (95.90) | 8.22 pg/mL (5.93) | $A^+$ (<0.001) $C^+$ (<0.001) | 4.4 / 5.5 |
| IL-13 | 0.89 pg/mL (1.69) | 1.66 pg/mL (2.39) | 56.50 pg/mL (12.77) | 35.39 pg/mL (14.71) | $A^+$ (<0.001) $B^+$ (<0.001) $C^+$ (<0.001) | 9.2 / 5.6 |
| IL-15 | 1.41 pg/mL (0.38) | 1.17 pg/mL (0.76) | 34.58 pg/mL (9.49) | 19.34 pg/mL (8.38) | $A^+$ (<0.001) $B^+$ (<0.001) $C^+$ (<0.001) | 2.0 / 10.5 |
| IL-18 | 795.44 pg/mL (454.99) | 534.89 pg/mL (380.24) | 1,250.97 pg/mL (489.78) | 825.03 pg/mL (476.15) | $A^+$ (<0.001) $B^+$ (<0.001) $C^+$ (<0.001) $D^-$ (<0.001) | 2.6 / 4.1 |
| IL-27 | 146.94 pg/mL (77.36) | 203.92 pg/mL (122.13) | 712.49 pg/mL (328.81) | 179.85 pg/mL (126.77) | $A^+$ (<0.001) $C^+$ (<0.001) $D^+$ (0.007) | 3.6 / 10.1 |
| IL-37 | 5.01 pg/mL (9.35) | 18.35 pg/mL (62.46) | 52.21 pg/mL (166.11) | 12.84 pg/mL (32.96) | $A^+$ (0.003) $C^+$ (<0.001) | 9.5 / 11.2 |
| IP-10 | 601.55 pg/mL (456.06) | 1002.60 pg/mL (3,398.17) | 41,744.72 pg/mL (24,057.54) | 16,328.46 pg/mL (10,563.07) | $A^+$ (<0.001) $B^+$ (<0.001) $C^+$ (<0.001) | 1.2 / 7.2 |

| | | | | | | |
|---|---|---|---|---|---|---|
| LBP | 5,044.63 ng/mL (1,877.45) | 2,784.96 ng/mL (1,294.61) | 2,703.05 ng/mL (1,192.72) | 2,763.58 ng/mL (1,227.00) | $D^-$ (<0.001) | 4.6 / 6.3 |
| Leptin | 38,154.39 pg/mL (43,582.59) | 36,826.33 pg/mL (45,789.10) | 34,285.98 pg/mL (38,489.85) | 31,536.79 pg/mL (36,972.26) | None | 6.2 / 9.1 |
| LIF | 7.17 pg/mL (54.91) | 4.42 pg/mL (11.97) | 6.44 pg/mL (9.00) | 11.20 pg/mL (11.08) | $B^+$ (<0.05) | 4.3 / 13.0 |
| MCP-1 | 369.53 pg/mL (189.10) | 1,362.45 pg/mL (2,294.33) | 27,108.60 pg/mL (30,915.41) | 251,479.46 pg/mL (125,104.65) | $A^+$ (0.002) $B^+$ (<0.001) $C^-$ (<0.001) | 7.0 / 8.7 |
| MIG | 98.80 pg/mL (54.16) | 79.05 pg/mL (67.80) | 370.60 pg/mL (438.56) | 469.69 pg/mL (560.99) | $A^+$ (<0.001) $B^+$ (<0.001) | 2.2 / 5.5 |
| MIP-1α | 17.56 pg/mL (12.78) | 154.94 pg/mL (399.23) | 203,194.27 pg/mL (87,288.06) | 17,418.50 pg/mL (20,265.90) | $A^+$ (<0.001) $B^+$ (0.004) $C^+$ (<0.001) | 4.1 / 24.0 |
| MIP-1β | 77.29 pg/mL (42.39) | 722.80 pg/mL (1,080.23) | 361,176.25 pg/mL (163,794.24) | 37,543.06 pg/mL (32,770.33) | $A^+$ (<0.001) $B^+$ (<0.001) $C^+$ (<0.001) | 2.1 / 10.2 |
| PAI-1 | 2.38 ng/mL (1.53) | 4.24 ng/mL (1.95) | 4.42 ng/mL (1.60) | 5.50 ng/mL (1.84) | $B^+$ (<0.001) $C^-$ (<0.001) $D^+$ (<0.001) | 5.2 / 5.4 |
| SAA | 8,652.40 ng/mL (22,128.75) | 3,689.80 ng/mL (10,828.99) | 3,727.61 ng/mL (11,118.24) | 3,610.03 ng/mL (9,877.76) | $D^-$ (<0.001) | 3.8 / 11.3 |
| sCD30 | 1,269.11 pg/mL (1,912.35) | 3,583.20 pg/mL (10,126.96) | 4,596.40 pg/mL (11,374.34) | 4,196.08 pg/mL (7,911.59) | $D^+$ (<0.001) | 9.0 / 27.5 |
| sICAM-1 | 525.62 ng/mL (139.71) | 320.10 ng/mL (97.66) | 328.62 ng/mL (87.26) | 338.01 ng/mL (93.77) | $B^+$ (0.008) $D^-$ (<0.001) | 3.8 / 9.9 |
| sVCAM-1 | 530.27 ng/mL (153.07) | 347.53 ng/mL (107.19) | 354.77 ng/mL (97.47) | 372.29 ng/mL (97.53) | $B^+$ (0.002) $D^-$ (<0.001) | 6.2 / 7.1 |
| TGF-α | 2.89 pg/mL (7.80) | 6.71 pg/mL (8.07) | 48.19 pg/mL (19.05) | 42.91 pg/mL (18.55) | $A^+$ (<0.001) $B^+$ (<0.001) $C^+$ (<0.001) $D^+$ (0.015) | 1.8 / 2.8 |
| TGF-β1 | 3,274.63 pg/mL (2,586.43) | 24,592.04 pg/mL (9,939.31) | 23,622.09 pg/mL (8,998.65) | 39,836.08 pg/mL (17,886.74) | $B^+$ (<0.001) $C^-$ (<0.001) $D^+$ (<0.001) | 11.1 / 9.8 |

| | | | | | | |
|---|---|---|---|---|---|---|
| TNF-α | 1.44 pg/mL (1.04) | 5.45 pg/mL (8.16) | 6,758.40 pg/mL (3,467.24) | 408.64 pg/mL (689.30) | $A^+$ (<0.001)<br>$C^+$ (<0.001) | 3.6 / 37.9 |
| TNFRI | 3,656.14 pg/mL (1,271.55) | 2,488.13 pg/mL (1,052.58) | 3,489.48 pg/mL (1,217.64) | 3,924.66 pg/mL (1,362.12) | $A^+$ (<0.001)<br>$B^+$ (<0.001)<br>$C^-$ (<0.001)<br>$D^-$ (<0.001) | 1.56 / 4.65 |
| TRAIL | 128.22 pg/mL (42.27) | 172.79 pg/mL (71.27) | 289.85 pg/mL (96.40) | 161.49 pg/mL (58.38) | $A^+$ (<0.001)<br>$C^+$ (<0.001)<br>$D^+$ (<0.001) | 1.7 / 16.7 |

The four numeric columns (Plasma, 24-hr Null, 24-hr FLU, 24-hr LPS) report mean concentrations with SD in parentheses. Values for 24-hr Null, FLU, and LPS samples were dilution-corrected using the urea method. For each biomarker, we performed a repeated-measures ANOVA (within-subject factor: sample type); Tukey's HSD post-hoc tests were conducted only when the omnibus ANOVA reached α = 0.05. The "Significant 24h inducer" column indicates which pairwise comparisons were significant and the direction of change: **A**=LPS vs. Null; **B**=FLU vs. Null; **C**=LPS vs. FLU; **D**=Null vs. Plasma. A plus sign **(+)** indicates the first condition was significantly higher; a minus sign **(–)** indicates it was significantly lower. Parenthetical values are Tukey HSD p-values (two-sided). "None" indicates ANOVA p < 0.05 but no post-hoc comparison survived correction; "NS" indicates ANOVA p > 0.05. Intra-/inter-assay %CVs are reported as "intra/inter." All TruCulture concentrations (Null, LPS, FLU) reflect urea-based dilution correction. For visualization, all six possible pairwise comparisons are displayed in the interactive Shiny application (https://primeknee.shinyapps.io/prime_viz/), which shows the full comparison set and underlying distributions for each biomarker. **Biomarker abbreviations**: Activin A (Activin A), CRP (C-reactive protein), D-Dimer (D-dimer), Eotaxin (CCL11), Fibrinogen (Fibrinogen), GDF-15 (Growth/differentiation factor 15), GRO-α (CXCL1), Granzyme B (Granzyme B), gp130 (IL-6 receptor subunit β), IFN-α2a (Interferon-α2a), IFN-γ (Interferon-γ), I-TAC (CXCL11), IL-1β, IL-1Ra, IL-2, IL-4, IL-5, IL-6, IL-7, IL-8 (CXCL8), IL-10, IL-12p70, IL-13, IL-15, IL-18, IL-27, IL-37, IP-10 (CXCL10), LBP, Leptin, LIF, MCP-1 (CCL2), MIG (CXCL9), MIP-1α (CCL3), MIP-1β (CCL4), PAI-1, SAA, sCD30, sICAM-1, sVCAM-1, TGF-α, TGF-β1, TNF-α, TNFRI, and TRAIL.

**Table S2A. Stably-selected Plasma and PBMC-stimulated biomarkers associated with Expected Recovery Differential (ERD) measures of resilience after knee surgery.**

| Expected Recovery Differential (ERD) | | | | | |
|---|---|---|---|---|---|
| **Sample Type** | **Biomarker Stability Classification** | **Pain Intensity Scores** | **Pain Interference Scores** | **Independence in Lower Extremity ADLs** | **Step Counts** |
| **PLASMA** | | | | | |
| Absolute concentration | Sample size | 129 | 130 | 130 | 101 |
| | Biomarkers with 10/10 stability score | **IL-12p70; IL-5; IFN-α2a; MIP-1β; CD30; LIF; LBP** | **IL-5; IL-8; IL-7; SAA; LBP; Activin A** | **IFN-y; IL-12p70; IL-13; eotaxin; CD30; leptin; TNFRI** | **IFN-y; IL-1β; IL-2; IL-37; TNFRI; D-dimer** |
| | Biomarkers with 1-9 stability score | MIP-1α; D-dimer; IL-2; MCP-1; IL-1β; GDF-15; TNFRI; IL-37; IL-10; IL-1Ra | CD30; TGF-α; LIF; D-dimer; IL-10; IL-15; MCP-1; IL-27; I-TAC; IL-12p70; IL-13; IL-1β; MIP-1β; TRAIL; sICAM-1 | IL-2; Activin A; MIG; TRAIL; MIP-1β; IL-37 | gp130; CD30; MIP-1β; TGF-α; IL-4; IL-6; TNF-α; IL-10; IL-5; MIP-1α |
| **LPS-Stimulated Biomarkers (dilution factor corrected)** | | | | | |
| **Sample Type** | **Biomarker Stability Classification** | **Pain Intensity Scores** | **Pain Interference Scores** | **Independence in Lower Extremity ADLs** | **Step Counts** |
| Absolute 24hr LPS STIM | Sample size | 139 | 137 | 142 | 110 |
| | Biomarkers with 10/10 stability score | **IL-37; silCAM-1; LBP; TGF-β1** | **IL-7; SAA; silCAM-1; LBP; PAI-1; Activin A** | **eotaxin; CD30; leptin; LIF; TNFRI; gp130; TGF-β1** | **IL-13; TNF-α; CD30; leptin; LIF; TNFRI; PAI-1** |
| | Biomarkers with 1-9 stability score | IL-18; GDF-15; D-dimer; IL-2; SAA; TNFRI; Activin A; IL-27; IL-15; IP-10; sVCAM-1; IL-1β; IL-8; eotaxin | IL-37; TNF-α; MIP-1β; LIF; MCP-1; TRAIL; TGF-α; D-dimer; TNFRI | IL-27; TRAIL; Granzyme B; IL-18; IL-7; IL-15; LBP | IL-7; IL-37; LBP; IP-10; I-TAC; IL-10; D-dimer |
| 24hr LPS STIM/NULL ratio | Sample size | 136 | 137 | 140 | 108 |
| | Biomarkers with 10/10 stability score | **IL-13; IL-2; IL-7; CD30; TNFRI** | **IL-10; IL-13; IFN-α2a; IL-15; MIP-1α; IL-37; LIF; TNFRI** | **IL-12p70; IL-13; IL-1β; IL-2; IL-5; IFN-α2a; IL-37** | **IL-10; IL-8; GRO-α; LIF; TGF-β1** |
| | Biomarkers with 1-9 stability score | TGF-β1; IL-12p70; IL-15; IP-10; MIP-1α; IL-1β; IL-27; IFN-y; fibrinogen; IL-37; IL-10 | Activin A; Granzyme B; IL-12p70; I-TAC | I-TAC; fibrinogen; Granzyme B; IL-27; IL-4; MIP-1β | IL-2; CD30; IL-5; MIP-1α; MIP-1β; MCP-1; IL-37; IFN-y; IL-6; MIG; TGF-α |
| %change [(24hr LPS STIM - NULL) / NULL] | Sample size | 136 | 137 | 140 | 108 |
| | Biomarkers with 10/10 stability score | **IL-12p70; IL-15; IL-7; IP-10; leptin; SAA; TNFRI** | **IL-13; IL-4; IL-15; SAA; LIF; TNFRI** | **IL-12p70; IL-13; IL-1β; IL-5; IFN-α2a; MIG; fibrinogen** | **IFN-y; IL-10; IL-8; LIF** |
| | Biomarkers with 1-9 stability score | IFN-y; IL-13; fibrinogen; IL-1Ra; IL-37; LBP; CD30 | IL-37; Activin A; IFN-α2a; IL-12p70; Granzyme B; LBP; TGF-β1 | I-TAC; IL-37; IL-27; Granzyme B; gp130; MIP-1β; IL-1Ra; TGF-β1 | IL-2; MIP-1α; PAI-1; IL-1β; GRO-α; MCP-1; eotaxin; IL-5; MIP-1β; CD30; IL-6 |
| **FLU-Stimulated Biomarkers (dilution factor corrected)** | | | | | |
| **Sample Type** | **Biomarker Stability Classification** | **Pain Intensity Scores** | **Pain Interference Scores** | **Independence in Lower Extremity ADLs** | **Step Counts** |
| | Sample size | 13 | 139 | 139 | 107 |

| | | | | | |
|---|---|---|---|---|---|
| Absolute 24hr FLU STIM | Biomarkers with 10/10 stability score | **IL-10; IL-6; TRAIL; GDF-15; silCAM-1; LIF; LBP** | **IL-10; Granzyme B; IL-37; SAA; silCAM-1; LIF; LBP** | **eotaxin; CD30; IL-37; leptin; TNFRI; gp130** | **IL-7; CD30; I-TAC; LIF; TNFRI; PAI-1; TGF-β1; D-dimer** |
| | Biomarkers with 1-9 stability score | CD30; IL-27; IL-1Ra; TNFRI; IP-10; TGF-β1; D-dimer | IL-4; I-TAC; IL-1β; IP-10; CD30; D-dimer; leptin; GDF-15; IL-27; TGF-β1 | TRAIL; IL-5; IL-2; IL-8; SAA; IL-12p70; IL-27; IL-15; sVCAM-1 | Granzyme B; LBP; IP-10; GRO-α; IL-27; Activin A |
| 24hr FLU STIM/NULL ratio | Sample size | 135 | 136 | 137 | 105 |
| | Biomarkers with 10/10 stability score | **IL-13; IL-2; IL-27; TGF-β1** | **IL-10; IL-13; IL-5; IFN-α2a; IL-27; IL-37; TRAIL** | **IL-13; IL-1β; IL-5; IFN-α2a; IP-10; SAA; fibrinogen** | **IL-10; CD30; GRO-α; IL-37; LIF; TGF-β1** |
| | Biomarkers with 1-9 stability score | IL-8; IL-15; IL-10; GRO-α; IL-12p70; IL-5; IL-6; IL-37; MIG; IFN-y; IL-1β; LIF; IP-10 | IP-10; IL-1Ra; IL-4; IL-1β; IL-2; IL-15; MIG; LIF; TNFRI | IL-6; CD30; PAI-1; Granzyme B; IL-4; MCP-1; MIP-1β; TNFRI | IL-4; IL-2; IL-8; MIP-1β; IL-6; MIP-1α; IL-15; TNF-α; IFN-y; IL-5; IFN-α2a |
| %change [(24hr FLU STIM - NULL) / NULL] | Sample size | 135 | 136 | 137 | 105 |
| | Biomarkers with 10/10 stability score | **IFN-y; IL-13; IL-8; IL-27; TGF-β1** | **IL-10; IL-13; IL-4; IFN-α2a; IL-27; TRAIL; TNFRI** | **IL-13; IL-1β; IL-5; IFN-α2a; IP-10** | **IL-10; GRO-α; LIF; TGF-β1** |
| | Biomarkers with 1-9 stability score | IL-15; IL-37; CRP; IL-12p70; TNFRI; IL-2; IL-5; GRO-α; D-dimer; IFN-α2a; I-TAC | IL-37; IL-15; IP-10; IL-1Ra; IFN-y; IL-5 | IL-12p70; SAA; TGF-β1; fibrinogen; MCP-1; MIG; silCAM-1; TNFRI; D-dimer | GDF-15; MIP-1α; D-dimer; IL-2; IL-8; TRAIL; IL-5; IL-37; IFN-α2a; MIP-1β; TNF-α; IL-1Ra; IFN-y; IL-1β; CD30 |

**Table S2B. Stably-selected Plasma and PBMC-stimulated biomarkers associated with Resilience Trajectory (RT) measures of resilience after knee surgery.**

| **Resilience Trajectory (RT)** | | | | | |
|---|---|---|---|---|---|
| **Sample Type** | **Biomarker Stability Classification** | **Pain Intensity Scores** | **Pain Interference Scores** | **Independence in Lower Extremity ADLs** | **Step Counts** |
| **PLASMA** | | | | | |
| Absolute concentration | Sample size | 150 | 150 | 150 | 113 |
| | Biomarkers with 10/10 stability score | **IL-12p70; IFN-α2a; IL-15; IL-37; MIG; SAA** | **IL-5; IL-7; IL-27; leptin; LBP** | **IL-10; IL-5; leptin; TNFRI; fibrinogen** | **IFN-y; IL-1β; IL-4; leptin; fibrinogen** |
| | Biomarkers with 1-9 stability score | IL-5; IL-27; LBP; Granzyme B; IL-1Ra; IL-1β; CD30; TRAIL | IL-1Ra; IL-37; LIF; IFN-α2a; IL-15; IFN-y; IL-1β; TNF-a; TNFRI | IL-8; MCP-1; CD30; IFN-y; eotaxin; IL-2; GDF-15; GRO-α; IL-13; IL-7 | MCP-1; TRAIL; silCAM-1; IL-27; IL-10; IL-6; gp130; GDF-15; MIP-1β; Granzyme B; IL-37; sVCAM-1 |
| **LPS Stimulated Biomarkers (dilution factor corrected)** | | | | | |
| **Sample Type** | **Biomarker Stability Classification** | **Pain Intensity Scores** | **Pain Interference Scores** | **Independence in Lower Extremity ADLs** | **Step Counts** |
| Absolute 24hr LPS STIM | Sample size | 162 | 162 | 162 | 123 |
| | Biomarkers with 10/10 stability score | **IL-2; IL-37; GDF-15; CRP; LBP** | **eotaxin; leptin; TNFRI; LBP** | **IL-7; MIP-1β; CD30; leptin; GDF-15; TNFRI** | **IL-12p70; IL-7; leptin; GDF-15; PAI-1** |
| | Biomarkers with 1-9 stability score | silCAM-1; IP-10; leptin; MIG; SAA; gp130; TNFRI; TGF-β1; I-TAC; IL-27; TGF-α; IL-10; IL-12p70; IFN-α2a; IL-15; fibrinogen | IL-7; LIF; IP-10; MCP-1; silCAM1; IL-37; TRAIL; IL-10; Granzyme B; IL-1Ra; IL-8; IL-27; SAA; PAI-1; D-dimer; fibrinogen | IP-10; LBP; IL-12p70; eotaxin; TGF-β1; TGF-α; LIF; fibrinogen; PAI-1 | TRAIL; IL-15; TNF-α; IL-10; CRP; LBP; TGF-β1; IL-18; IL-5; IL-8; CD30; GRO-α; I-TAC |
| 24hr LPS STIM/NULL ratio | Sample size | 159 | 159 | 159 | 121 |
| | Biomarkers with 10/10 stability score | **IL-13; CD30; TRAIL; LIF; PAI-1** | **IL-15; MIP-1α; IL-27; IL-37; LIF** | **IFN-y; IL-4; IL-5; IL-6; LIF; PAI-1; TGF-β1** | **IL-2; IL-4; TNF-α; GRO-a; TGF-α** |
| | Biomarkers with 1-9 stability score | IL-15; IL-1Ra; TNFRI; D-dimer; GRO-α; Granzyme B; Activin A; MIP-1α; IL-12p70; IL-4; IP-10 | IFN-α2a; SAA; IL-13; CD30; TNFRI; Activin A; IP-10; IL-2; TRAIL; PAI-1; IL-5; TGF-β1; fibrinogen | IL-13; IFN-α2a; TRAIL; eotaxin; IL-2; D-dimer; IL-15; GRO-α; Granzyme B; IL-27 | IL-5; LIF; IL-10; TGF-β1; MIG; IL1-5; CD30; IL-1Ra; D-dimer; IL-12p70; IL-6; IFN-α2a; MIP-1β |
| %change [(24hr LPS STIM - NULL) / NULL] | Sample size | 159 | 159 | 159 | 121 |
| | Biomarkers with 10/10 stability score | **IL-13; CD30; leptin; LIF** | **IL-5; IL-15; MIP-1α; IL-37; LIF** | **IL-4; IL-5; IL-6; LIF; TGF-β1** | **IL-2; IL-4; GRO-a** |
| | Biomarkers with 1-9 stability score | IL-15; D-dimer; IL-5; PAI-1; TNFRI; GRO-α; TRAIL; IL-4; IL-1Ra; fibrinogen; MIP-1α; CRP; silCAM-1; Activin A | CD30; TNFRI; IFN-α2a; TGF-β1; IFN-y; fibrinogen; IL-4; MCP-1; IL-27; IP-10; Activin A | IFN-y; IFN-α2a; PAI-1; D-dimer; IL-15; eotaxin; TRAIL; IL-27; GRO-α; IL-1β | IL-5; TNF-α; leptin; eotaxin; IL-18; IL-10; IL-8; IL-6; CD30; gp130; IFN-α2a; IL-13; IL-1β; MCP-1; MIP-1α; |

| | | | | | |
|---|---|---|---|---|---|
| | | | | | MIP-1β; LIF; TGF-α |

| FLU-Stimulated Biomarkers (dilution factor corrected) | | | | | |
|---|---|---|---|---|---|
| **Sample Type** | **Biomarker Stability Classification** | **Pain Intensity Scores** | **Pain Interference Scores** | **Independence in Lower Extremity ADLs** | **Step Counts** |
| Absolute 24hr FLU STIM | Sample size | 159 | 159 | 159 | 120 |
| | Biomarkers with 10/10 stability score | **IL-10; IL-37; TRAIL; LIF; LBP** | **eotaxin; IL-27; leptin; LIF; TNFRI; LBP** | **IL-15; leptin; GDF-15; TNFRI; LBP** | **IL-5; leptin; GDF-15; PAI-1** |
| | Biomarkers with 1-9 stability score | D-dimer; siICAM-1; PAI-1; SAA; IL-2; GDF-15; eotaxin; sVCAM-1; IL-7; IP-10; TNFRI | IL-10; IL-37; siICAM-1; PAI-1; IP-10; IL-6; TGF-β1 | IL-12p70; IL-1β; IL-5; eotaxin; IL-2; CD30; gp130; PAI-1; IFN-α2a; TGF-β1; IL-27; IL-37 | TGF-β1; CD30; TRAIL; LIF; I-TAC; siICAM-1; IL-7; IL-37; LBP; MCP-1; GRO-α; IL-27; sVCAM-1 |
| 24hr FLU STIM/NULL ratio | Sample size | 156 | 156 | 156 | 118 |
| | Biomarkers with 10/10 stability score | **IL-13; IL-8; IL-15; GRO-α; Granzyme B; IL-27; LIF; Activin A** | **IL-12p70; IL-13; IL-15; IL-1Ra; IL-37; LIF; Activin a** | **IL-4; IL-5; IL-15; IP-10; IL-27; LIF** | **IL-2; IL-4; IL-5; IL-6; CD30; GRO-α; LIF; TGF-β1** |
| | Biomarkers with 1-9 stability score | TNF-α; eotaxin; TGF-β1; IP-10; IL-37 | IL-27; MIG; IL-4; MIP-1α; IFN-α2a; eotaxin; IL-1β | IL-13; TGF-β1; IFN-y; PAI-1; IL-10; IL-12p70; IL-1β; IL-37 | MIG; IL-1β; IFN-α2a; IL-37; MIP-1α; I-TAC |
| %change [(24hr FLU STIM - NULL) / NULL] | Sample size | 156 | 156 | 156 | 118 |
| | Biomarkers with 10/10 stability score | **IL-13; IL-15; GROa; Granzyme B; IL-27; Activin A** | **IL-12p70; IL-15; IL1Ra; MIG; LIF** | **IL-4; IL-5; IP-10; PAI-1; TGF-β1** | **IL-2; IL-4; IL-5; IL-6; GRO-α; LIF** |
| | Biomarkers with 1-9 stability score | IL-8; CRP; TRAIL; LIF; eotaxin; IL-7; IFN-y; IL-4; IP-10; TNF-α | Activin A; IFN-y; IL-4; IL-5; CRP; GDF-15; IL-8; eotaxin; IL-27; IL-37; IL-13; IL-6; IP-10 | IL-10; IL-15; IL-27; LIF; IL-1Ra; IFN-y; leptin; fibrinogen; TRAIL; IL-1β; GRO-α | fibrinogen; IL-1β; CD30; I-TAC; SAA; TNF-α; TGF-β1 |

The table lists all biomarkers selected by LASSO for the interim stable set in at least one of ten separate analytic configurations (using all combinations of 2 sampling methods and 5 penalty parameters) as predictors of resilience outcomes (**S2A**) ERD and (**S2B**) RT. Biomarkers in bold font were selected all 10 times/iterations. LASSO selection was performed on biomarkers from traditional plasma and stimulated whole blood whose values were defined in one of three manners: 1) absolute biomarker concentrations of the stimulated samples; 2) stim/null ratios, defined as the ratio of the 24-hour stimulated value to its paired 24-hour unstimulated control; and 3) percent (% (x/x%) change, defined as (24-hour stimulated minus 24-hour unstimulated)/24-hour unstimulated. Prior to LASSO selection, all stimulated biomarker concentrations were corrected for the dilution factors inherent in the TruCulture tube collection system, using the urea method. Color coding is based on unadjusted beta estimates of biomarkers and outcomes: **red** positive association, **black** negative association; The sample size represents number of individuals in the specific analysis.

**Table S3. Frequency and percentage of biomarker selection by resilience outcome (ERD, RT, ERD+RT) across ALL measures (LPS, FLU and plasma in bold black font) and for plasma (PL in red font) alone.**

| Biomarker | ERD ALL n/% (PL n/%) | RT ALL n/% (PL n/%) | ERD & RT ALL n/% (PL n/%) | Biomarker | ERD ALL n/% (PL n/%) | RT ALL n/% (PL n/%) | ERD & RT ALL n/% (PL n/%) |
|---|---|---|---|---|---|---|---|
| LBP | 73/61% (20/50%) | 84/70% (19/48%) | 157/65% (39/49%) | IFN-γ | 20/17% (20/50%) | 21/18% (21/53%) | 41/17% (41/51%) |
| Leptin | 42/35% (10/25%) | 95/79% (30/75%) | 137/57% (40/50%) | IP-10 | 15/13% (0/0%) | 25/21% (0/0%) | 40/17% (0/0%) |
| TNFRI | 75/63% (23/58%) | 56/47% (11/28%) | 131/55% (34/43%) | MCP-1 | 11/9% (7/18%) | 26/22% (18/45%) | 37/15% (25/31%) |
| CD30 | 89/74% (36/90%) | 32/27% (10/25%) | 121/50% (46/58%) | Activin A | 35/29% (19/48%) | 0/0% (0/0%) | 35/15% (19/24%) |
| LIF | 70/58% (15/38%) | 49/41% (9/23%) | 119/50% (24/30%) | MIP-1β | 23/19% (18/45%) | 12/10% (2/5%) | 35/15% (20/25%) |
| IL-37 | 58/48% (13/33%) | 59/49% (20/50%) | 117/49% (33/41%) | Granzyme B | 24/20% (0/0%) | 10 8% (7/18%) | 34/14% (7/9%) |
| GDF-15 | 24/20% (3/8%) | 62/52% (7/18%) | 86/36% (10/13%) | IFN-α2a | 10/8% (10/25%) | 21/18% (18/45%) | 31/13% (28/35%) |
| IL-7 | 41/34% (10/25%) | 45/38% (11/28%) | 86/36% (21/26%) | I-TAC | 19/16% (2/5%) | 9/8% (0/0%) | 28/12% (2/3%) |
| sICAM-1 | 41/34% (1/3%) | 45/38% (9/23%) | 86/36% (10/13%) | IL-8 | 16/13% (10/25%) | 12/10% (9/23%) | 28/12% (19/24%) |
| IL-5 | 29/24% (21/53%) | 49/41% (29/73%) | 78/33% (50/63%) | TNF-α | 19/16% (3/8%) | 8/7% (1/3%) | 27/11% (4/5%) |
| TRAIL | 34/28% (3/8%) | 42/35% (10/25%) | 76/32% (13/16%) | Fibrinogen | 0/0% (0/0%) | 25/21% (20/50%) | 25/10% (20/25%) |
| Eotaxin | 31/26% (10/25%) | 40/33% (5/13%) | 71/30% (15/19%) | IL-1RA | 8/7% (1/3%) | 17/14% (14/35%) | 25/10% (15/19%) |
| IL-27 | 30/25% (3/8%) | 41/34% (25/63%) | 71/30% (28/35%) | MIG | 8/7% (8/20%) | 15/13% (10/25%) | 23/10% (18/23%) |
| IL-10 | 26/22% (5/13%) | 44/37% (15/38%) | 70/29% (20/25%) | IL-13 | 21/18% (11/28%) | 1/1% (1/3%) | 22/9% (12/15%) |
| PAI-1 | 30/25% (0/0%) | 39/33% (0/0%) | 69/29% (0/0%) | TGF-α | 15/13% (12/30%) | 6/5% (0/0%) | 21/9% (12/15%) |
| SAA | 41/33% (10/25%) | 23/19% (10/25%) | 64/27% (20/25%) | IL-4 | 10/8% (4/10%) | 10/8% (10/25%) | 20/8% (14/18%) |
| IL-12p70 | 25/21% (21/53%) | 35/29% (10/25%) | 60/25% (31/39%) | IL-6 | 14/12% (4/10%) | 6/5% (4/10%) | 20/8% (8/10%) |
| IL-2 | 34/28% (23/58%) | 23/19% (4/10%) | 57/24% (27/34%) | CRP | 0/0% (0/0%) | 16/13% (0/0%) | 16/7% (0/0%) |
| TGF-β1 | 32/27% (0/0%) | 24/20% (0/0%) | 56/23% (0/0%) | IL-18 | 9/8% (0/0%) | 3/3% (0/0%) | 12/5% (0/0%) |
| D-Dimer | 46/38% (19/48%) | 9/8% (0/0%) | 55/23% (19/24%) | GRO-α | 2/2% (0/0%) | 6/5% (3/8%) | 8/3% (3/4%) |
| IL-15 | 8/7% (3/8%) | 37/31% (18/45%) | 45/19% (21/26%) | sVCAM-1 | 3/3% (0/0%) | 4/3% (1/3%) | 7/3% (1/1%) |
| IL-1β | 22/18% (14/35%) | 22/18% (13/33%) | 44/18% (27/34%) | MIP-1α | 6/5% (6/15%) | 0/0% (0/0%) | 6/3% (6/8%) |
| gp130 | 29/24% (9/23%) | 12/10% (4/10%) | 41/17% (13/16%) | | | | |

Ranked by overall frequency of selection. ERD and RT equating to 240 possible chances of selection overall (*ALL from plasma, LPS or FLU stimulated) or 80 possible chances of selection in plasma (PL). ERD=Expected Recovery Differential; RT=Resilience Trajectory

**Table S4. The twenty-nine clinical predictors used to construct the expected recovery differential (ERD) outcome.**

| Variable | Description |
|---|---|
| Age at Surgery | Continuous (in years) |
| Gender | Categorical (Male / Female) |
| Race | Categorical (White / Black / Asian / More than One / Missing) |
| Ethnicity | Categorical (Not Hispanic / Hispanic / Missing) |
| Financial Stress | Categorical (Enough / Little to Spare / Need to Cut Back / Have Difficulty / Missing) |
| Education | Continuous (in years completed) if data available, plus a missing indicator |
| Chronic Lung Disease | Categorical (Yes / No / Missing) |
| Vascular Disease | Categorical (Cardiac / Peripheral / CV / No / Missing) |
| Heart Disease | Categorical (Preserved / Reduced / Yes Unspecified / No / Missing) |
| Chronic Liver Disease | Categorical (Yes / No / Missing) |
| Diabetes | Categorical (Type I, Type II, Yes Unspecified, No, Missing) |
| History of treated non-skin cancer | Categorical (Breast / Prostate / Hematopoetic / Other / Yes Unspecified / No / Missing) |
| eGFR | Continuous if data available, plus a missing indicator |
| BMI | Continuous (in kg/m^2) |
| 3-minute walk | Continuous (in feet) |
| grip strength | Continuous (in kg) |
| 3MS | Continuous if data available, plus a missing indicator |
| Trail Making Test B | Continuous (in sec) if data available, plus a missing indicator |
| Trail Making Test A | Continuous (in sec) if data available, plus a missing indicator |
| 15 item word list | Continuous (number of items recalled) if data available, plus a missing indicator |
| Digit Symbol Substitution Test | Continuous (number completed) if data available, plus a missing indicator |
| Resilience Scale | Continuous if data available, plus a missing indicator |
| PHQ-9 | Continuous if data available, plus a missing indicator |
| PROMIS SF Emotional Support 4a | Continuous (t-score) if data available, plus a missing indicator |
| Admission Patient Class during Surgery | Categorical (Inpatient / Outpatient) |
| Anesthesia Type During Surgery | Categorical (General / MAC / Spinal Regional) |
| Estimated Blood Loss During Surgery | Continuous if data available, plus a missing indicator |
| Anesthesia Time During Surgery | Continuous (in hours) |
| Baseline Outcome (e.g., Pain Intensity for the Pain Intensity ERD) | Continuous if data available, plus a missing indicator |

All measures derived from baseline visit except for age derived at the time of surgery.
